# A novel classifier of radiographic knee osteoarthritis for use on knee DXA images is predictive of joint replacement in UK Biobank

**DOI:** 10.1101/2024.08.08.24311666

**Authors:** Rhona A Beynon, Fiona R Saunders, Raja Ebsim, Benjamin G Faber, Mijin Jung, Jennifer S Gregory, Claudia Lindner, Richard M Aspden, Nicholas C Harvey, Timothy Cootes, Jonathan H Tobias

## Abstract

**Objectives:** DXA scans may offer a novel means of evaluating radiographic knee osteoarthritis (rKOA) in large population studies and through opportunistic screening. We aimed to develop and apply a semi-automated method for assessing rKOA using ∼20,000 knee DXA images from UK Biobank (UKB) and assess its face validity by checking for expected relationships with clinical outcomes.

**Methods:** Right knee DXA scans were manually annotated for osteophytes to derive corresponding grades. Joint space narrowing (JSN) grades in the medial joint compartment were determined from automatically measured minimum joint space width. Overall rKOA grade (0-4) was determined by combining osteophyte and JSN grades. Logistic regression was employed to investigate the associations of osteophyte, JSN, and rKOA grades with knee pain and hospital-diagnosed knee osteoarthritis (HES-KOA). Cox proportional hazards modelling was used to examine the associations of these variables with risk of subsequent total knee replacement (TKR).

**Results:** Of the 19,595 participants included (mean age: 63.7), 19.5% had rKOA grade ≥1 (26.1% female; 12.5% male). Grade ≥1 osteophytes and grade ≥1 JSN were associated with knee pain, HES-KOA, and TKR. Higher rKOA grades were linked to stronger associations with these clinical outcomes, with the most pronounced effects observed for TKR. HRs for the association of rKOA grades with TKR were 3.28, 8.75, and 28.63 for grades 1, 2 and 3-4, respectively.

**Conclusions:** Our DXA-derived measure of rKOA demonstrated a progressive relationship with clinical outcomes. These findings support the use of DXA for classifying rKOA in large epidemiological studies and in future population-based screening.

**Key messages:**

- Radiographic knee osteoarthritis (rKOA) can be semi-automatically derived from DXA images.
- DXA-derived rKOA shows expected relationships with clinical outcomes of knee osteoarthritis.
- DXA imaging presents a viable method for classifying rKOA in large-scale epidemiological research.

## Introduction

Knee osteoarthritis (KOA) is the most common form of osteoarthritis, affecting 5.4 million people in the UK alone (1). Annually, this results in around 100,000 knee replacements being performed (2), with demand for these procedures expected to rise by nearly 40% by 2060 (3). Diagnosis of KOA is primarily based on clinical symptoms, with persistent knee pain being the most common. Radiographically, KOA displays distinctive features such as osteophyte formation, joint space narrowing (JSN), subchondral sclerosis and cysts. These features have been integrated into grading systems for use in epidemiological studies, including the widely used Kellgren-Lawrence (KL) grading system (4), which classifies KOA severity into five grades, ranging from 0 for “normal” up to 4 for “severe”. Typically, a KL score of ≥2, indicating the presence of a definite osteophyte and possible JSN, is used to define radiographic knee osteoarthritis (rKOA) in research studies (5). Applying this approach to large epidemiological studies is challenging due to its time-consuming and subjective nature (5-9). However, there is growing interest in developing computer-aided techniques to enhance reliability and reduce the time required to derive these grades. (10-15).

To date, large scale epidemiological studies of osteoarthritis have primarily been based on plain radiographs (X-rays), for which KL scoring was initially developed. Dual-energy X-ray Absorptiometry (DXA) imaging has recently emerged as a viable alternative (16). Initially developed for measuring bone mineral density (BMD) at the hip and spine, DXA is widely used as a tool for diagnosing osteoporosis. Advancements in DXA technology have significantly improved resolution, enabling the visualisation of features such as osteophytes and measurements of JSN (16). The very low radiation exposure associated with these devices (17) make them highly suitable for use in large scale epidemiological studies, as well as offering potential application in population screening.

UK Biobank (UKB), a large prospective cohort study, is acquiring hip and knee DXA images from 100,000 participants (18). A recent proof-of-concept study, involving 40,000 of these hip DXA scans, suggested DXA images can be used for accurately classifying hip osteoarthritis; robust associations were observed between grades 2-4 of radiographic hip osteoarthritis (rHOA) and different clinical outcomes, including a nearly 60-fold greater likelihood of requiring total hip replacement in individuals with rHOA grade 4 (19). While similar investigations for the knee are lacking, a prior study based on knee DXA scans in UKB suggested that a DXA-derived imaging biomarker for knee shape, derived from a statistical shape model, could predict the need for total knee replacement (TKR) (20).

The primary objectives of this study were to develop a semi-automated method for classifying rKOA using DXA scans, to apply this method to a large dataset of images from UKB, and to evaluate its face validity by examining its relationship with clinically important KOA outcomes.

## Methods

### Population

This study included participants from the UKB Extended Imaging Study, a subset of the larger UKB cohort. UKB enrolled ∼500,000 participants aged 40-69 from across the UK between 2006 and 2010, collecting extensive health and lifestyle data. The Extended Imaging Study, initiated in 2014, aimed to collect medical imaging data, including DXA scans, from 100,000 participants (18). UKB has full ethical approval from the National Information Governance Board for Health and Social Care and the North-West Multi-Centre Research Ethics Committee (11/NW/0382). All UKB participants provided consent, including permission for their health to be followed-up through linkage to health-related records. This study was approved by UKB under application number 17295.

### DXA-based measures of knee osteoarthritis

#### DXA-based scoring of osteophytes and joint space narrowing

High resolution knee DXA-scans were acquired using a Lunar iDXA scanner (GE-Healthcare, Madison, WI, USA), with participants lying in a supine position. A machine-learning algorithm based on random-forest regression voting (BoneFinder®, The University of Manchester (21)), initially trained on ∼7000 manually annotated left knee DXA images, placed 129 points along the bone contours of the distal femur, proximal tibia, proximal fibula, and superior patella, excluding osteophytes. Details of this methodology have been published previously (20). The present study is based on a selection of ∼20,000 randomly selected right knee DXA images with automated point placement checked by trained annotators (RB, FS).

At the time point placement was checked, each image was also evaluated for the presence of medial and lateral femoral and tibial osteophytes. If osteophytes were present, they were shaded manually (**Figure 1**), and the osteophyte area (mm^2^) was calculated using a custom tool (University of Manchester). Osteophytes were then automatically graded on a scale of 0-3 based on area thresholds derived from manual grading (**supplementary methods**).

**Figure 1:**
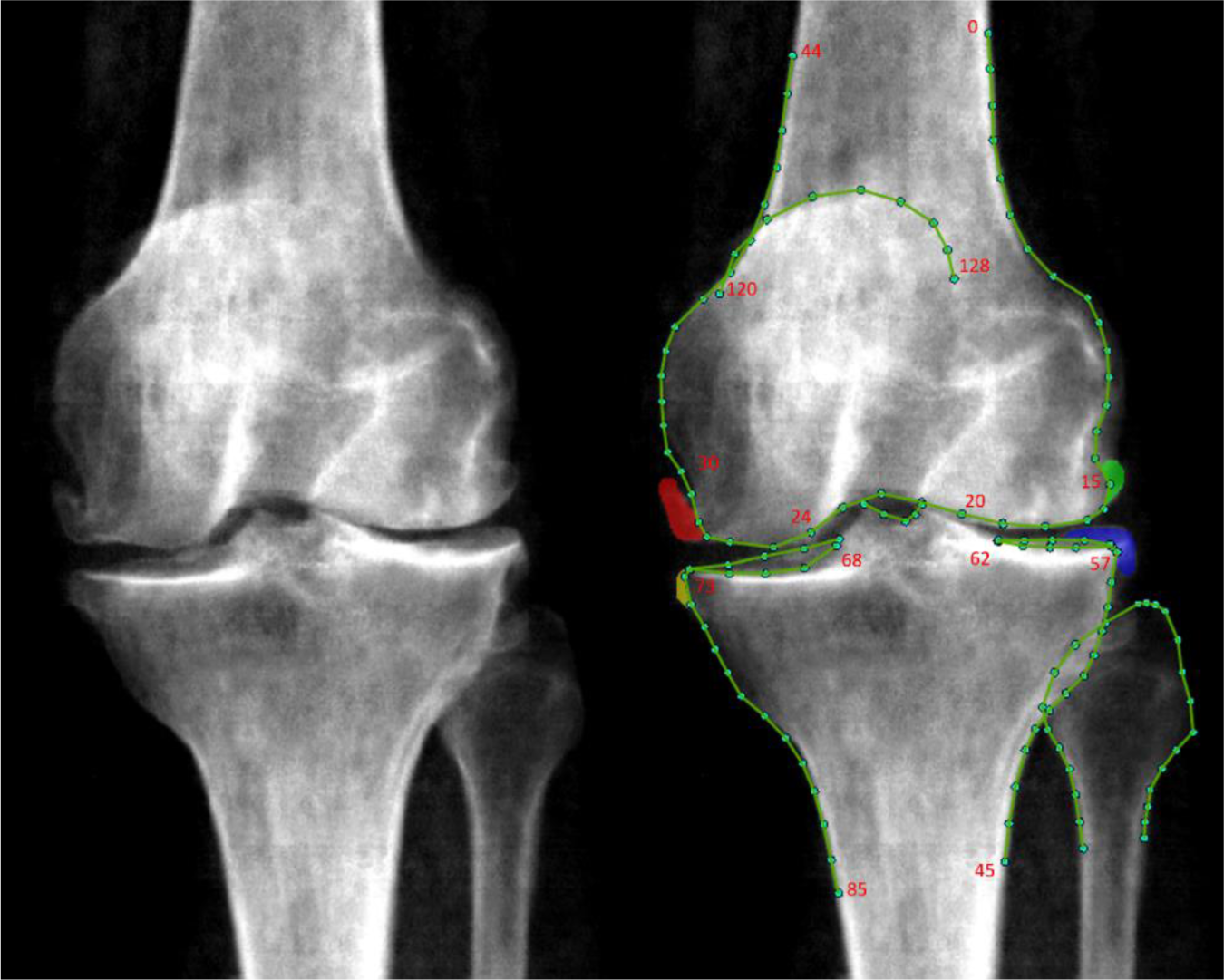
An example DXA scan with osteophytes marked up. The left panel displays a DXA image of the knee without annotations, while the right panel shows the same DXA image with osteophytes manually shaded. Osteophytes are indicated by colours corresponding to their locations: red for the medial femur, green for the lateral femur, yellow for the medial tibia, and blue for the lateral tibia. Minimum joint space width (mJSW) was measured at specific points in the medial and lateral compartments. For the distal femur, mJSW was measured between medial points 24-30 and lateral points 15-20. For the proximal tibia, mJSW was measured between medial points 68-73 and lateral points 57-62.

Minimum Joint Space Width (mJSW; mm) of the medial joint compartment was automatically measured between predefined points (**Figure 1**) using a custom Python 3.0 script. Medial joint space narrowing (JSN) grades were assigned based on the mJSW measurements: JSN grade 0 for mJSW >3mm; grade 1 for mJSW >2.5mm and ≤3mm; grade 2 for mJSW >2mm and ≤2.5mm; grade 3 for mJSW <2mm. The medial joint compartment was selected due to its common involvement in primary KOA, with preliminary analyses indicating it as the most reliable predictor of clinical outcomes.

#### Generation of rKOA grades

Overall rKOA grades were determined by integrating osteophyte and JSN grades. Subchondral sclerosis and cysts were not considered because they were rarely observed. Four osteophyte sites were assessed, each graded on a scale of 0 to 3, resulting in a total possible score of 12. To adjust for their relative contribution, each site’s grade was multiplied by 0.5, resulting in a maximum combined osteophyte score of 6. This score was then added to the JSN total, resulting in a maximum sum score of 9 (see supplementary Table 1). To provide a five-point overall rKOA grade (similar to the KL radiograph grading system), we used the following cut-offs: rKOA grade 0, sum score = 0; grade 1 >0 & ≤1.5; grade 2 >1.5 & ≤3; grade 3 >3 & ≤4.5; grade 4 >4.5. Figure 2 illustrates an example image corresponding to each rKOA grade. Additionally, as a sensitivity analysis, we adjusted mJSW measurements by normalising them against the mean height of the population before assigning JSN grades (**Supplementary Methods**).

**Figure 2:**
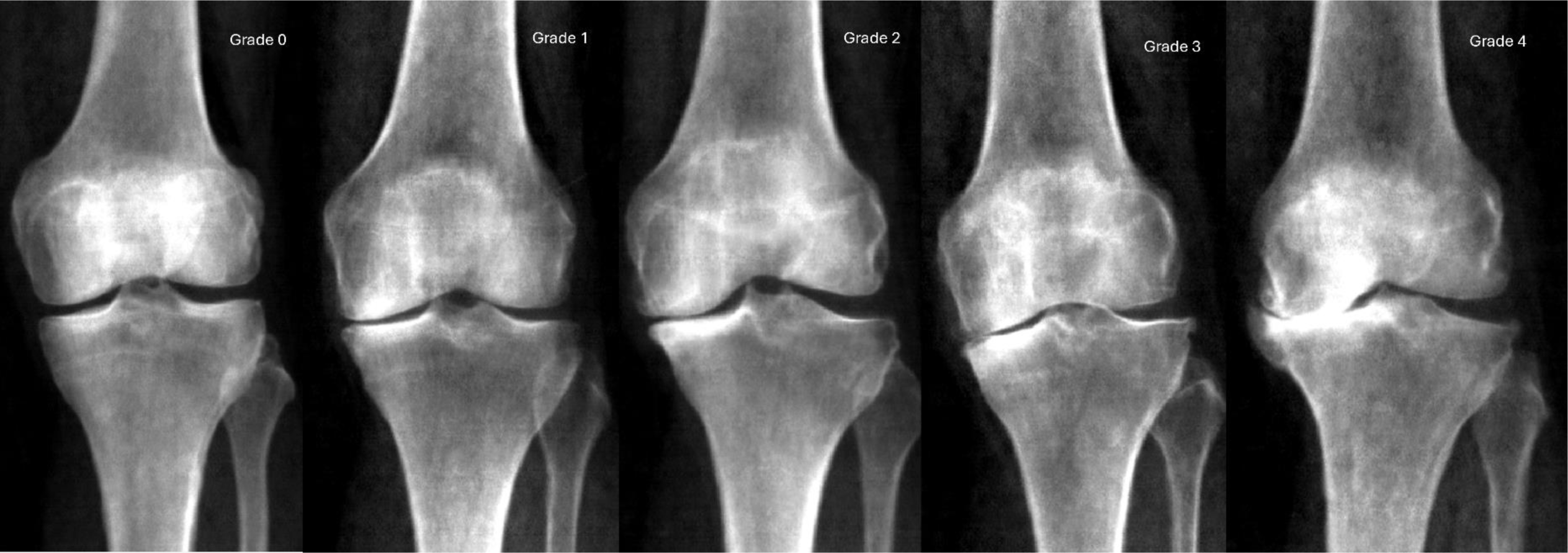
Example DXA scans representing each grade of radiographic knee osteoarthritis. Radiographic knee osteoarthritis (rKOA) grades were generated by integrating data on osteophyte grades and joint space narrowing (JSN) grades. The images show progression from Grade 0 to Grade 4, demonstrating increasing severity of osteoarthritic changes. Grade 0 indicates no radiographic features of osteoarthritis, while Grades 1 to 4 show progressively more significant joint space narrowing and osteophyte formation.

### Clinical outcomes

A binary variable indicating knee pain lasting for more than three months was created based on responses obtained from a questionnaire administered during the participants’ DXA appointment. Hospital-diagnosed KOA, hereafter referred to as HES-KOA, was determined using International Classification of Diseases (ICD) codes (9^th^ and 10^th^ revisions), which were obtained via linkage to Hospital Episodes Statistics (HES). Records began in 1997 and data were downloaded in July 2023, capturing information up until the end of October 2022. This variable was analysed cross-sectionally, recognising that KOA is a chronic condition that could have been present before the diagnosis. TKR had to be subsequent to the scan date and was based on Office of Population Censuses and Surveys (OPCS) codes, for which an associated date was obtained. None of the three clinical outcomes were side-specific.

### Statistical analysis

Logistic regression was employed to investigate the associations between osteophytes, JSN, and rKOA grades with knee pain and HES-KOA. The findings are presented as odds ratios (ORs) alongside their corresponding 95% confidence intervals (CIs). When assessing the association of these exposures with TKR, Cox proportional hazards modelling was used, with results reported as hazard ratios (HRs) along with their 95% CIs. The proportional hazards assumption was checked using Schoenfeld residuals. Each exposure was compared against a reference group of individuals with a grade of 0 for that specific exposure. Both crude and adjusted models were conducted, with adjustments made for age, sex, height, weight, and ethnicity (**Supplementary Methods**). The primary analysis included both males and females, with additional separate analyses conducted for each sex. Additionally, an interaction term for sex was incorporated into the primary models. All analyses were conducted using Stata version 17 (StataCorp, College Station, TX, USA).

## Results

### Population Characteristics

In total, 19,595 right knee DXA scans were available after applying quality control measures. The mean age of participants was 63.7 years (range 45 to 82 years), with approximately equal sex distribution (51.8 females) (Table 1). A total of 2,886 (14.7%) reported having had knee pain for >3 months, 917 (4.7%) had HES-KOA and 271 (1.4%) had undergone TKR subsequent to their DXA scan. The median time to TKR was 2.6 years (IQR: 1.3 to 4.1 years).

**Table 1:** Baseline descriptive statistics of the study population.

|  | All<br>(n=19,595) | Female<br>(n=10,146) | Male<br>(n=9449) |
| --- | --- | --- | --- |
| **Demographics** |  |  |  |
|  |  | *Mean (Range)* |  |
| Age (years) | 63.73 (45, 82) | 63.03 (45, 82) | 64.49 (45, 82) |
| Height (cm) | 170.20 (135, 202) | 163.59 (135, 198) | 177.29 (150, 202) |
| Weight (kg) | 75.27 (36, 169) | 67.96 (36, 169) | 83.12 (48, 160) |
| Ethnic background |  | *Frequency (%)* |  |
| White | 18963 (96.77%) | 9827 (96.86%) | 9136 (96.69%) |
| Asian | 263 (1.34%) | 124 (1.22%) | 139 (1.47%) |
| Chinese | 54 (0.28%) | 36 (0.35%) | 18 (0.19%) |
| Black | 116 (0.59%) | 63 (0.62%) | 53 (0.56%) |
| Mixed | 91 (0.46%) | 49 (0.48%) | 42 (0.44%) |
| Other | 110 (0.56%) | 61 (0.60%) | 49 (0.52%) |
| Unknown | 52 (0.27%) | 22 (0.22%) | 30 (0.32%) |
| **Radiographic measures** |  |  |  |
|  |  | *Frequency (%)* |  |
| OP any location | 2359 (12.04%) | 1473 (14.52%) | 886 (9.38%) |
| OP all locations | 71 (0.36%) | 43 (0.42%) | 28 (0.30%) |
| Medial femoral OP | 1328 (6.78%) | 900 (8.87%) | 428 (4.53%) |
| Lateral femoral OP | 303 (1.55%) | 194 (1.91%) | 109 (1.15%) |
| Medial tibial OP | 1003 (5.12%) | 596 (5.87%) | 407 (4.31%) |
| Lateral tibial OP | 1243 (6.34%) | 748 (7.37%) | 495 (5.24%) |
| medial JSN | 1847 (9.43%) | 1427 (14.06%) | 420 (4.44%) |
|  |  | *Mean (Range)* |  |
| Total OP area (mm<sup>2</sup>) | 24.28 (1.99, 316.62) | 22.88 (1.99, 316.62) | 26.59 (2.12, 266.69) |
| Medial femoral OP area (mm<sup>2</sup>) | 18.29 (1.99, 142.71) | 17.28 (1.99, 142.71) | 20.42 (3.95, 123.22) |
| Lateral femoral OP area (mm<sup>2</sup>) | 20.72 (2.46, 155.11) | 17.26 (2.46, 68.56) | 26.90 (3.29, 155.11) |
| Medial tibial OP area (mm<sup>2</sup>) | 11.57 (2.05, 118.09) | 10.56 (2.05, 118.09) | 13.06 (2.12, 95.09) |
| Lateral tibial OP area (mm<sup>2</sup>) | 12.14 (2.18, 121.00) | 11.38 (2.19, 121.00) | 13.28 (2.18, 90.43) |
| Medial mJSW (mm) | 3.90 (0, 7.27) | 3.61 (0, 6.31) | 4.21 (0.37, 7.27) |
| Lateral mJSW (mm) | 4.15 (0.47, 7.87) | 3.73 (0.48, 7.74) | 4.59 (0.47, 7.87) |
| **Clinical outcomes** |  |  |  |
|  |  | *Frequency (%)* |  |
| Knee pain >3 months | 2886 (14.73%) | 1500 (14.78%) | 1386 (14.67%) |
| HES-KOA | 917 (4.68%) | 425 (4.19%) | 492 (5.21%) |
| TKR | 271 (1.38%) | 141 (1.39%) | 130 (1.38%) |
|  |  | *Median (IQR)* |  |
| Time to TKR (years) | 2.60 (1.31, 4.10) | 2.54 (1.46, 3.96) | 2.68 (1.31, 4.62) |

Abbreviations: HES-KOA, Knee Osteoarthritis Based on Hospital Episodes Statistics; mJSW, Minimum Joint Space Width; OP, Osteophyte; TKR, Total Knee Replacement.

### Prevalence of rKOA

#### Osteophytes and JSN

Osteophytes (grade ≥1) were detected in 2,359 (12.0%) DXA scans (Table 1), with the greatest prevalence observed in the medial femur, followed by the lateral tibia, medial tibia, and lateral femur. Notably, females exhibited a higher frequency of osteophytes across all sites compared to males. Osteophytes tended to be larger on the femur than on the tibia. Medial JSN (grade ≥1) was present in 1,847 participants (9.4%) and was almost three times more common in females. The prevalence of individual osteophyte and JSN grades can be found in Supplementary Table 2 and Supplementary Table 3, respectively.

#### Overall rKOA grade

A classifier for rKOA was constructed by combining scores for osteophytes and JSN. The distribution of participants across different rKOA grades is detailed in Supplementary Table 4, with participant characteristics categorised by rKOA grade presented in Supplementary Table 5. Among the participants, 15,768 (80.5%) exhibited grade 0 rKOA, while 2,883 (14.7%) had grade 1, 712 (3.6%) had grade 2, 158 (0.8%) had grade 3 and 74 (0.4%) had grade 4. Due to the small number of participants with KOA grade 4, rKOA grades 3 and 4 were combined in subsequent analyses. Among females, 26.1% had an rKOA grade 1 or higher, compared to 12.5% for males.

### Associations between rKOA and knee osteoarthritis outcomes

#### Osteophytes versus knee osteoarthritis outcomes

In adjusted analyses, the presence of one or more osteophyte (grade ≥1) at any site was associated with knee pain, HES-KOA and TKR, with progressively higher effect estimates (OR 3.38 [95% CI: 3.06, 3.75], 4.67 [4.03, 5.42] and 7.51 [5.84, 9.66], respectively) (Table 2). Results from the unadjusted analysis, provided in Supplementary Table 6, demonstrated similar associations, albeit with larger effect sizes. Osteophytes located at each knee region were related to all clinical outcomes, with lateral femoral and medial tibial osteophytes showing the strongest associations with HES-KOA and TKR. Results of the sex-stratified analysis are detailed in Supplementary Tables 7 and 8. The point estimates for the associations of individual osteophyte sites (grade ≥1) with pain and TKR were higher in males than in females after adjustment. However, no evidence of a sex interaction was observed when an interaction term was included in the main model.

**Table 2:** The adjusted associations of osteophytes and joint space narrowing (grades ≥1) with knee osteoarthritis outcomes.

|  | Pain |  |  | HES-KOA |  |  | TKR |  |  |
| --- | --- | --- | --- | --- | --- | --- | --- | --- | --- |
|  | OR | 95% CI | p-value | OR | 95% CI | p-value | HR | 95% CI | p-value |
| Any OP | 3.38 | (3.74, 0.00) | $1.0 \times 10^{-123}$ | 4.67 | (4.03, 5.42) | $2.7 \times 10^{-93}$ | 7.51 | (5.84, 9.66) | $8.4 \times 10^{-56}$ |
| OP at all locations | 3.90 | (6.30, 0.00) | $3.1 \times 10^{-08}$ | 4.37 | (2.48, 7.68) | $3.2 \times 10^{-07}$ | 5.77 | (2.94, 11.31) | $3.4 \times 10^{-07}$ |
| Medial femoral OP | 3.50 | (3.96, 0.00) | $1.2 \times 10^{-87}$ | 3.84 | (3.23, 4.57) | $8.5 \times 10^{-52}$ | 5.20 | (3.98, 6.80) | $2.6 \times 10^{-33}$ |
| Lateral femoral OP | 3.39 | (4.30, 0.00) | $1.7 \times 10^{-23}$ | 6.19 | (4.71, 8.14) | $4.1 \times 10^{-39}$ | 7.93 | (5.60, 11.23) | $2.0 \times 10^{-31}$ |
| Medial tibial OP | 4.17 | (4.77, 0.00) | $2.8 \times 10^{-93}$ | 5.43 | (4.55, 6.47) | $9.4 \times 10^{-39}$ | 8.06 | (6.21, 10.46) | $1.9 \times 10^{-55}$ |
| Lateral tibial OP | 2.90 | (3.30, 0.00) | $3.0 \times 10^{-59}$ | 3.65 | (3.06, 4.36) | $6.1 \times 10^{-47}$ | 4.86 | (3.72, 6.35) | $5.8 \times 10^{-31}$ |
| JSN | 1.45 | (1.65, 0.00) | $7.0 \times 10^{-09*}$ | 2.23 | (1.85, 2.67) | $1.0 \times 10^{-17*}$ | 3.23 | (2.45, 4.26) | $8.0 \times 10^{-17*}$ |

Logistic regression and cox proportional hazards modelling results showing the associations of osteophyte and JSN grades (grade $\geq 1$ vs 0) with knee pain, hospital diagnosed knee osteoarthritis and knee replacement, respectively (n=19,595). Models were adjusted for age, sex, height, weight and ethnicity. Abbreviations: CI, Confidence Interval; HES-KOA, Knee Osteoarthritis Based on Hospital Episodes Statistics; HR, Hazard Ratio; JSN, Joint Space Narrowing; OP, Osteophyte; OR Odds Ratio; TKR, Total Knee Replacement. \*Denotes a sex-interaction term with $p < 0.05$ .

Higher osteophyte grades were generally more strongly associated with the three clinical outcomes at all four sites, in both unadjusted and adjusted analyses (Figure 3; tabulated in Supplementary Tables 9 (unadjusted) and 10 (adjusted)). However, some exceptions were observed: grade 2 medial tibial osteophytes showed slightly weaker associations with pain compared with grade 1 osteophytes; grade 3 lateral tibial osteophytes showed slightly weaker associations with HES-KOA compared with grade 2 osteophytes; grade 2 lateral femoral osteophytes showed slightly weaker associations with TKR compared with grade 1 osteophytes. Although the trend was less evident in the sex-stratified analyses, (Supplementary Tables 11-14), the ORs and HRs for grade 3 osteophytes were generally larger than for grade 1. There was no evidence that sex modified the associations of osteophyte grades with clinical outcomes (Supplementary Tables 9 and 10).

**Figure 3:**
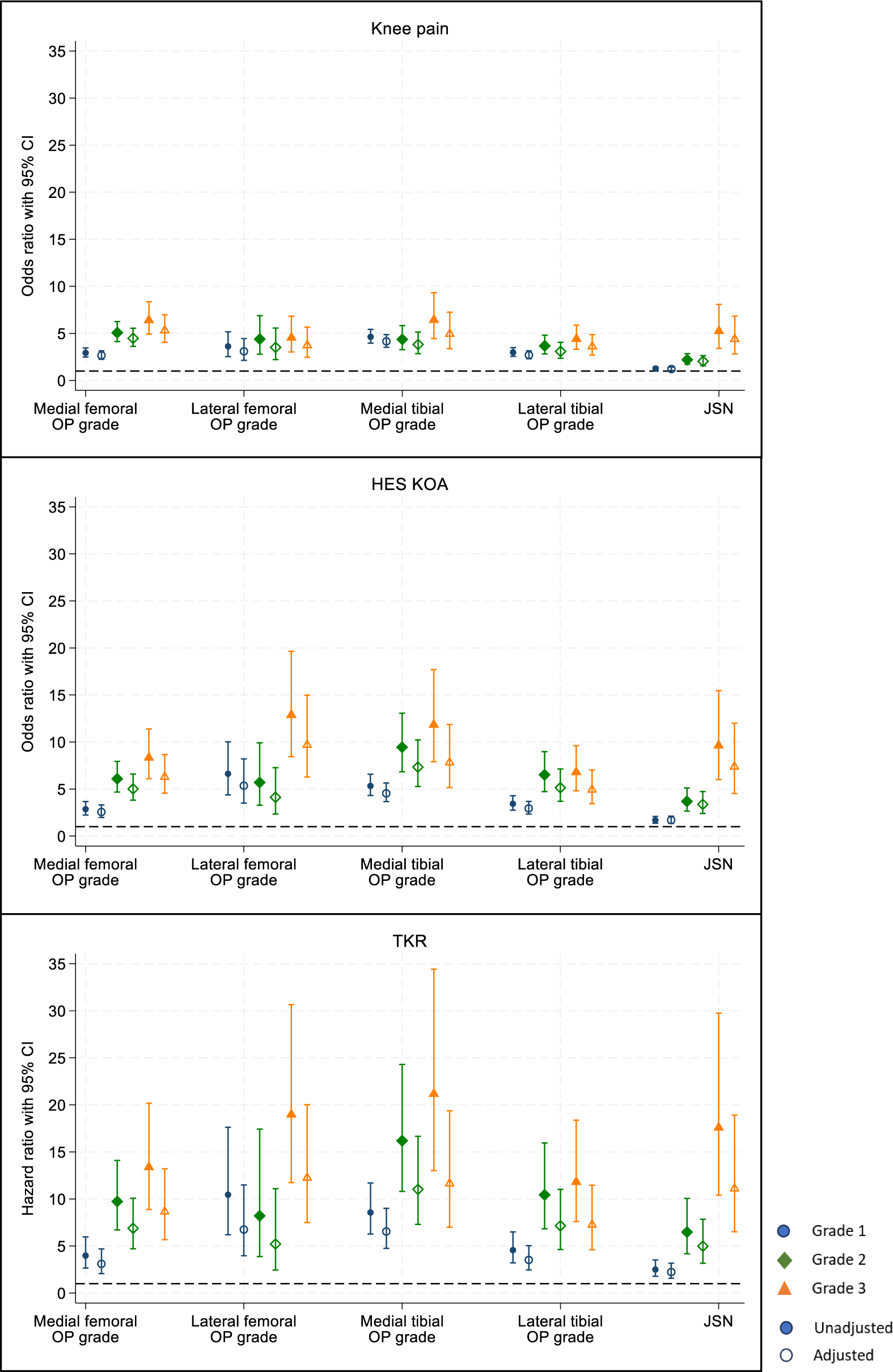
Associations of Osteophyte grades and medial joint space narrowing grades with knee osteoarthritis outcomes. The graphs depict both crude and adjusted odds ratios and hazard ratios, accompanied by 95% confidence intervals, for knee pain, HES-KOA, and TKR, across different grades of osteophytes and JSN (n=19,595). Models are adjusted for age, sex, height, weight, and ethnicity. Abbreviations: CI, Confidence Interval; HES-KOA, Knee Osteoarthritis based on Hospital Episode Statistics; JSN, Joint Space Narrowing; OP, Osteophyte; TKR, Total Knee Replacement.

#### JSN versus knee osteoarthritis outcomes

JSN (grade ≥1) was associated with all three clinical outcomes, with effect sizes almost 50% less than that of osteophytes (knee pain: adjusted OR 1.45 [1.28,1.65]; HES-KOA: OR 2.23 [1.85, 2.67] and TKR: HR 3.23 [2.45, 4.26]) (Table 2). In sex stratified analyses, ORs and HRs for the association of JSN (grade ≥1) with clinical outcomes were found to be higher in males compared with females (Supplementary Tables 7 and 8) and there was evidence of a sex interaction for all outcomes (Table 2).. As JSN grades increased, there was a corresponding increase in effect estimates (Figure 3), which stronger associations for HES-KOA compared with pain, and for TKR compared with HES-KOA. This trend remained consistent among females (Supplementary Tables 13 and 14) but was less evident in males (Supplementary Tables 11 and 12), though individual grades were still strongly associated with all three KOA outcomes. There was some evidence of a sex-interaction (9 and 10).

#### rKOA versus knee OA outcomes

The relationship between rKOA grades and clinical outcomes showed a consistent pattern of increasing strength of association with greater rKOA grade, which was observed across all three outcomes (Figure 4; tabulated in Supplementary Table 15). Adjusted ORs for pain ranged from 2.04 (1.84, 2.26) for grade 1 rKOA to 7.08 (5.41, 9.27) for grades 3-4. Similarly, for HES-OA, the adjusted ORs varied from 2.67 (2.25, 3.16) for grade 1 rKOA to 10.24 (7.53, 13.93) for grades 3-4. Regarding TKR, HRs ranged from 3.97 (2.90, 5.42) for grade 1 to 21.11 (14.28, 31.19) for grades 3-4. In sex-stratified analyses (Supplementary Table 15), rKOA measures remained associated with all three outcomes, with a clear progressive trend in females. In males, grade 2 rKOA had a stronger association with HES-KOA and TKR compared to grades 3-4, although grades 3-4 were still associated. Sex interactions were apparent for certain rKOA grades. Specifically, in adjusted models, there was evidence suggesting that the associations of rKOA grade 2 and rKOA grades 3-4 with HES-KOA and TKR were modified by sex. We conducted a sensitivity analysis to ensure that the observed associations between rKOA grades and clinical outcomes were not confounded by variations in participant height, as the shorter stature in females may have explained their greater prevalence of JSN. After deriving rKOA grades using height-normalised mJSW (prevalence detailed in Supplementary Table 16), the results showed a similar sex interaction in the relationship between rKOA grade and clinical outcomes (Supplementary Table 17).

**Figure 4:**
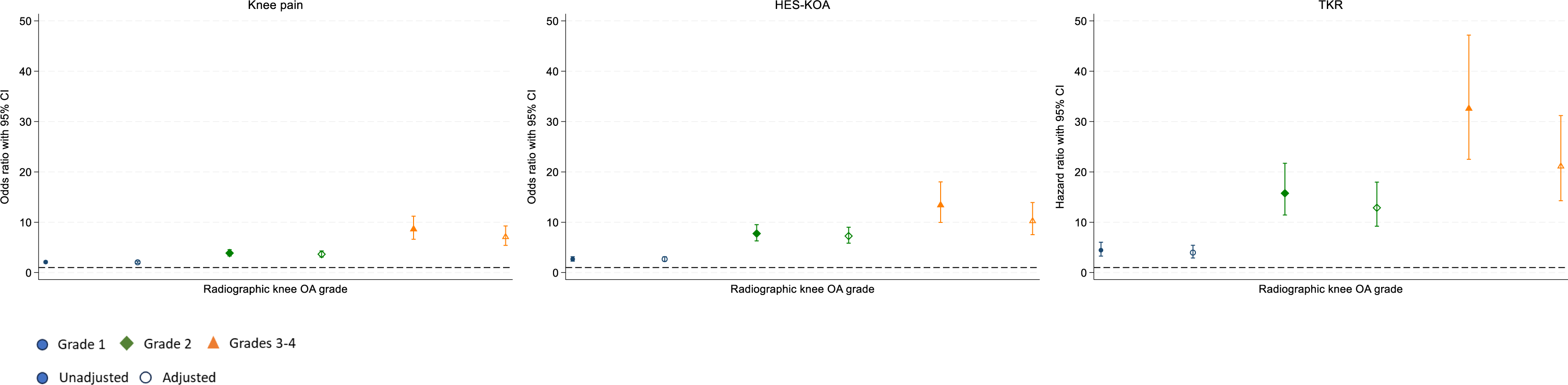
Associations of rKOA grade with knee osteoarthritis outcomes. The plots present both unadjusted and adjusted associations between rKOA grade, derived from a composite measure of osteophyte and JSN grades, and knee osteoarthritis outcomes (n=19,595). The models include adjustments for age, sex, height, weight, and ethnicity, with 95% confidence intervals provided. Abbreviations: CI, Confidence Interval; HES-KOA, Knee Osteoarthritis based on Hospital Episode Statistics; JSN, Joint Space Narrowing; rKOA, Radiographic Knee Osteoarthritis; TKR, Total Knee Replacement.

## Discussion

We aimed to create a novel classifier for rKOA based on knee DXA scans. Using semi-automated techniques to minimise subjective interpretation, we annotated and graded osteophytes and JSN on knee DXA scans from nearly 20,000 UKB participants to derive rKOA grade. Our study revealed an overall rKOA prevalence of 19.5%, consistent with previous estimates of rKOA based on plain radiographs (X-rays), though reported ranges vary widely (22). These variations likely stem from differences in participant selection, demographics, and the specific characteristics of the study populations. For instance, Cui et al. reported prevalence rates of rKOA (defined as KL ≥2) ranging from 9% to 55% across 19 studies conducted between 2001 to 2020, with a pooled estimate of 28.7% (23). All of these studies were notably smaller in scale compared to the present investigation. Specifically, the study reporting a 9% prevalence rate included 1,128 individuals from the USA with a mean age of 62 (range 34-90), whereas the study with a 55% prevalence rate involved 3,040 Japanese participants with a mean age of 70 (SD: 11). Our finding that women exhibited a higher rate than men (26.1% vs. 12.5%) aligns with the existing literature, which consistently shows a higher prevalence of KOA among women, especially over the age of 40 years (24, 25).

To evaluate the face validity of our measure of rKOA, we investigated associations with clinical outcomes related to KOA, namely prolonged knee pain, HES-KOA, and subsequent TKR. These outcomes serve as proxies for increasing severity, with TKR representing end-stage osteoarthritis. We observed robust and progressively increasing associations between grades of rKOA and all three outcomes. Furthermore, rKOA grades demonstrated stronger relationships with more advanced outcomes, with approximately seven-fold, ten-fold, and nineteen-fold increased risks of knee pain, HES-KOA, and TKR, respectively, for individuals classified with rKOA grades 3-4 compared to those with grade 0. These relationships appeared to reflect associations of both osteophyte and JSN grade with clinical outcomes, both of which were used to derived rKOA grade. That said, the presence of pain correlated more strongly with the presence of osteophytes than with JSN, which is consistent with some studies (26, 27), but not all (28). Taken together, these findings suggest that rKOA may have clinical relevance, given its relationship with outcomes such as pain and risk of TKR. While NICE guidelines prioritise symptom-based diagnosis (29), our results suggest that imaging could be beneficial in certain cases, potentially complementing clinical assessments and aiding in treatment decisions. Moreover, our proposed method for evaluating rKOA offers a means of evaluating structural changes associated with KOA in large cohorts. This could in turn to provide a basis for identifying new risk factors, including genetic factors, which could lead to the discovery of novel therapeutic targets.

Interestingly, we found evidence of a sex difference in the associations of JSN and rKOA with KOA outcomes, with generally higher effect estimates observed in males in sex-stratified analyses. This discrepancy may be attributed to narrower joint space width in healthy females compared with males, possibly due to their smaller stature, making JSN (derived from mJSW) a less specific measure for osteoarthritis in females. This could in turn result in weaker associations with clinical outcomes. However, results of the sensitivity analysis show that these differences persist even after normalising mJSW by mean height, suggesting that radiographic evidence of KOA may not correspond as closely with clinical outcomes in women. Other studies support this by demonstrating that, given the same level of radiographic severity, women tend to experience more intense pain and physical limitations than men (25, 30).

Similar to the conventional KL grading system, our DXA-derived classification system prioritises the assessment of osteophytes and JSN, as these features are considered hallmark signs of osteoarthritis progression and have been associated with knee symptoms (31-35). While sclerosis is a component of the KL grading system, definitive sclerosis was observed too infrequently on DXA images for inclusion in our classifier. This may represent a significant limitation given that some studies suggest sclerosis is associated with knee pain (28, 36). On the other hand, our classifier offers several distinct advantages. For example, KL grading often introduces ambiguity with terms like “definite” osteophyte and “possible” JSN, (assuming a continuous progression of these structural changes), which can lead to discrepancies between raters and across studies (5-9). In contrast, we automated the measurement of mJSW, enabling us to establish quantitative cut-offs for JSN. While osteophytes were manually identified, our classifier uses specific area-based cut-offs to define osteophyte grade, thereby avoiding the subjective scoring used in KL grading. Importantly, we observed strong correlations between these osteophyte grades and all three KOA outcomes, validating their use in future studies. By leveraging advancements in computer vision technologies, it may become feasible to fully automate the shading of osteophytes, enabling the widespread application of our classifier in large-scale epidemiological studies.

To our knowledge, no other automated or semi-automated classification system for rKOA on knee DXA images has been described previously. That said, various machine learning and deep-learning methods have been reported to analyse rKOA on X-rays. Rather than developing a classification system based on thresholds for JSN and osteophyte size, as here, X-ray based studies have generally trained models on diagnoses made by radiologists. For example, Thomas et al. developed an automated model to detect the presence of rKOA, defined as KL grade ≥ 2, using X-rays previously graded by a committee of radiologists (10). Their model, employing a convolutional neural network, demonstrated performance similar to that of radiologists. Similarly, Tiulpin et al. utilized deep-learning methods to accurately predict KL grade from knee X-rays, achieving an area under the ROC curve of 0.98 for detecting rKOA (KL ≥2) (37).

In terms of limitations, although our clinical outcomes related to KOA were not side-specific, our classification of rKOA was based solely on right knees. However, this approach likely reduces effect estimates rather than introducing biased associations. Additionally, HES-KOA, while specific, may be insensitive since obtaining an ICD code necessitates a hospital admission. Furthermore, since our classification system was developed using data from UKB, future studies should replicate our findings to validate their generalizability. A further limitation is that, unlike previous studies based on X-rays, DXA images are acquired with participants in a supine position, as opposed to weight bearing, meaning the mJSW is usually larger (38). Like X-rays, being two-dimensional, DXA scans provide a limited view of osteophytes and can be distorted by minor changes in patient positioning, potentially obscuring osteophytes from view. The lower prevalence of lateral femoral osteophytes observed in our analysis compared to other sites may indicate potential issues related to rotation during image acquisition.

In conclusion, we have developed a semi-automated classifier for rKOA for use on knee DXA images, based on combinations of osteophytes at four locations within the knee joint, and medial JSN. Having applied this classifier to right knee DXA images from ∼20,000 UKB participants, we observed expected prevalence rates for rKOA, including higher rates in females than males. Moreover, rKOA showed expected progressive associations with clinical outcomes namely knee pain, HES-OA and TKR. Based on these findings, we propose that knee DXA scans can provide a valuable tool in ascertaining rKOA in large cohort studies, as well as pointing to their possible use in population-based screening.

## Acknowledgements

The authors would like to thank the participants of the UK Biobank.

## Data availability

The data from this study will be available from UK Biobank in an upcoming data release. To access these resources, users must register with UK Biobank at: https://www.ukbiobank.ac.uk/enable-your-research/register. The BoneFinder® knee module and Markup-Tool are freely available on request: https://bone-finder.com/

## Supplementary Tables

**Supplementary Table 1:** The breakdown of the combine score including osteophyte and JSN grades. Radiographic osteophyte grades, initially multiplied by 0.5, were summed across the four sites on the medial and lateral aspects of the femur and tibia, with a maximum possible score of 6. This score was then combined with the JSN grade, resulting in a maximum possible score of 9 for each individual. Cut-offs were used to assign overall rKOA grades based on these combined scores.

| Combined score | N | % |
| --- | --- | --- |
| 0 | 15,768 | 80.47 |
| 0.5 | 823 | 4.2 |
| 1 | 1,678 | 8.56 |
| 1.5 | 382 | 1.95 |
| 2 | 428 | 2.18 |
| 2.5 | 142 | 0.72 |
| 3 | 142 | 0.72 |
| 3.5 | 71 | 0.36 |
| 4 | 55 | 0.28 |
| 4.5 | 32 | 0.16 |
| 5 | 29 | 0.15 |
| 5.5 | 17 | 0.09 |
| 6 | 11 | 0.06 |
| 6.5 | 7 | 0.04 |
| 7 | 3 | 0.02 |
| 7.5 | 5 | 0.03 |
| 8 | 1 | 0.01 |
| 9 | 1 | 0.01 |

**Supplementary Table 2:** Prevalence of Osteophytes by site.

| **Osteophyte grade** | **All**<br>(n=19,595) |  | **Female**<br>(n=10,146) |  | **Male**<br>(n=9,449) |  |
| --- | --- | --- | --- | --- | --- | --- |
|  | N | % | N | % | N | % |
| **Medial Femur** |  |  |  |  |  |  |
| 0 | 18,267 | (93.22) | 9,246 | (91.13) | 9,021 | (95.47) |
| 1 | 732 | (3.74) | 535 | (5.27) | 197 | (2.08) |
| 2 | 374 | (1.91) | 234 | (2.31) | 140 | (1.48) |
| 3 | 222 | (1.13) | 131 | (1.29) | 91 | (0.96) |
| **Lateral femur** |  |  |  |  |  |  |
| 0 | 19,292 | (98.45) | 9,952 | (98.09) | 9,340 | (98.85) |
| 1 | 130 | (0.66) | 100 | (0.99) | 30 | (0.32) |
| 2 | 78 | (0.40) | 49 | (0.48) | 29 | (0.31) |
| 3 | 95 | (0.48) | 45 | (0.44) | 50 | (0.53) |
| **Medial tibia** |  |  |  |  |  |  |
| 0 | 18,592 | (94.88) | 9,550 | (94.13) | 9,042 | (95.69) |
| 1 | 695 | (3.55) | 453 | (4.46) | 242 | (2.56) |
| 2 | 195 | (1.00) | 90 | (0.89) | 105 | (1.11) |
| 3 | 113 | (0.58) | 53 | (0.52) | 60 | (0.63) |
| **Lateral tibia** |  |  |  |  |  |  |
| 0 | 18,352 | (93.66) | 9,398 | (92.63) | 8,954 | (94.76) |
| 1 | 809 | (4.13) | 502 | (4.95) | 307 | (3.25) |
| 2 | 237 | (1.21) | 141 | (1.39) | 96 | (1.02) |
| 3 | 197 | (1.01) | 105 | (1.03) | 92 | (0.97) |

**Supplementary Table 3:** Prevalence of medial joint space narrowing (JSN).

|  | **All**<br>(n=19,595) | **Female**<br>(n=10,146) | **Male**<br>(n=9,449) |
| --- | --- | --- | --- |
| **Medial JSN grade** |  |  |  |
| 0 | 17748 (90.57%) | 8719 (85.94%) | 9029 (95.56%) |
| 1 | 1448 (7.39%) | 1129 (11.13%) | 319 (3.38%) |
| 2 | 315 (1.61%) | 248 (2.44%) | 67 (0.71%) |
| 3 | 84 (0.43%) | 50 (0.49%) | 34 (0.36%) |

**Supplementary Table 4:** Prevalence of radiographic knee osteoarthritis (rKOA).

|  | **All**<br>(n=19,595) | **Female**<br>(n=10,146) | **Male**<br>(n=9,449) |
| --- | --- | --- | --- |
| **rKOA grade** |  |  |  |
| 0 | 15768 (80.47%) | 7500 (73.92%) | 8268 (87.50%) |
| 1 | 2883 (14.71%) | 2027 (19.98%) | 856 (9.06%) |
| 2 | 712 (3.63%) | 492 (4.85%) | 220 (2.33%) |
| 3 | 158 (0.81%) | 87 (0.86%) | 71 (0.75%) |
| 4 | 74 (0.38%) | 40 (0.39%) | 34 (0.36%) |

**Supplementary Table 5:** Participant characteristics by radiographic KOA grade. Abbreviations: cm, centimetres; HES-KOA, hospital diagnosed knee osteoarthritis; JSN, joint space narrowing; Kg, kilograms; OP, osteophytes; TKR, total knee replacement.

|  | rKOA grade |  |  |  |
| --- | --- | --- | --- | --- |
|  | 1<br>N=2883 | 2<br>N=712 | 3<br>N=158 | 4<br>N=74 |
| **Demographics** | Mean (SD) |  |  |  |
| Age (years) | 65.22 (7.00) | 66.44 (6.89) | 66.66 (6.38) | 66.76 (6.13) |
| Height (cm) | 167.27 (9.08) | 167.28 (9.48) | 169.84 (9.70) | 168.69 (9.99) |
| Weight (kg) | 74.24 (15.76) | 76.39 (16.63) | 82.61 (17.49) | 90.00 (21.88) |
| **Radiographic measures** | Frequency (%) |  |  |  |
| OP at any site | 1668 (57.86%) | 459 (64.47%) | 158 (100.00%) | 74 (100.00%) |
| OP at all sites | 0 (0.00%) | 15 (2.11%) | 29 (18.35%) | 27 (36.49%) |
| Medial femoral OP | 805 (27.92%) | 326 (45.79%) | 130 (82.28%) | 67 (90.54%) |
| Lateral femoral OP | 89 (3.09%) | 114 (16.01%) | 66 (41.77%) | 34 (45.95%) |
| Medial tibial OP | 518 (17.97%) | 286 (40.17%) | 129 (81.65%) | 70 (94.59%) |
| Lateral tibial OP | 773 (26.81%) | 294 (41.29%) | 115 (72.78%) | 61 (82.43%) |
| Medial JSN | 1312 (45.51%) | 393 (55.20%) | 82 (51.90%) | 60 (81.08%) |
| **Clinical outcomes** | Frequency (%) |  |  |  |
| Knee pain | 635 (22.03%) | 245 (34.41%) | 83 (52.53%) | 42 (56.76%) |
| HES-KOA | 225 (7.80%) | 140 (19.66%) | 45 (28.48%) | 24 (32.43%) |
| Knee replacement | 75 (2.60%) | 62 (8.71%) | 25 (15.82%) | 15 (20.27%) |

**Supplementary Table 6:** The unadjusted associations of osteophytes and joint space narrowing (grades >1) with knee OA outcomes in males and females combined. Abbreviations: CI, Confidence Interval; HES-KOA, Knee osteoarthritis based on Hospital Episodes Statistics; HR, Hazard Ratio; JSN, Joint Space Narrowing; OP, Osteophyte; OR Odds Ratio; TKR, Total Knee Replacement. *Denotes a sex-interaction term with p<0.05. n=19,595.

|  | Pain |  |  | HES-KOA |  |  | TKR |  |  |
| --- | --- | --- | --- | --- | --- | --- | --- | --- | --- |
|  | OR | 95% CI | *p* | OR | 95% CI | *p* | HR | 95% CI | *p* |
| Any OP | 3.72 | (3.37, 4.10) | $<1.0 \times 10^{-40}$ | 5.39 | (4.69, 6.21) | $<1.0 \times 10^{-40}$ | 9.74 | (7.67, 12.38) | $<1.0 \times 10^{-40}$ |
| OP at all locations | 5.07 | (3.18, 8.10) | $1.04 \times 10^{-11}$ | 6.51 | (3.76, 11.28) | $2.26 \times 10^{-11}$ | 10.52 | (5.41, 20.44) | $3.93 \times 10^{-12}$ |
| Medial femoral OP | 3.95 | (3.50, 4.45) | $<1.0 \times 10^{-40}$ | 4.53 | (3.84, 5.35) | $<1.0 \times 10^{-40}$ | 7.10 | (5.50, 9.18) | $<1.0 \times 10^{-40}$ |
| Lateral femoral OP | 4.09 | (3.24, 5.16) | $2.04 \times 10^{-32}$ | 8.05 | (6.19, 10.49) | $<1.0 \times 10^{-40}$ | 12.36 | (8.83, 17.29) | $<1.0 \times 10^{-40}$ |
| Medial tibial OP | 4.75 | (4.16, 5.43) | $<1.0 \times 10^{-40}$ | 6.70 | (5.65, 7.94) | $<1.0 \times 10^{-40}$ | 11.32 | (8.82, 14.53) | $<1.0 \times 10^{-40}$ |
| Lateral tibial OP | 3.31 | (2.93, 3.76) | $<1.0 \times 10^{-40}$ | 4.48 | (3.77, 5.31) | $<1.0 \times 10^{-40}$ | 6.76 | (5.22, 8.75) | $<1.0 \times 10^{-40}$ |
| JSN | 1.53 | (1.35, 1.73) | $7.19 \times 10^{-12*}$ | 2.29 | (1.92, 2.72) | $1.26 \times 10^{-20*}$ | 3.80 | (2.92, 4.95) | $3.85 \times 10^{-23*}$ |

**Supplementary Table 7:** The associations of osteophytes and joint space narrowing (grades >1) with knee OA outcomes in males. Abbreviations: CI, Confidence Interval; HES-KOA, Knee osteoarthritis based on Hospital Episodes Statistics; HR, Hazard Ratio; JSN, Joint Space Narrowing; OP, Osteophyte; OR Odds Ratio; TKR, Total Knee Replacement. Models were adjusted for age, height, weight and ethnic group. n=9,449.

|  | Pain |  |  | HES-KOA |  |  | TKR |  |  |
| --- | --- | --- | --- | --- | --- | --- | --- | --- | --- |
|  | OR | 95% CI | *p* | OR | 95% CI | *p* | HR | 95% CI | *p* |
| Any OP | 3.72 | (3.20, 4.33) | $<1.0 \times 10^{-40}$ | 5.12 | (4.17, 6.28) | $<1.0 \times 10^{-40}$ | 10.45 | (7.41, 14.74) | $<1.0 \times 10^{-40}$ |
| OP at all locations | 5.87 | (2.79, 12.33) | $3.05 \times 10^{-06}$ | 6.14 | (2.60, 14.52) | $3.54 \times 10^{-05}$ | 11.06 | (4.09, 29.92) | $2.24 \times 10^{-06}$ |
| Medial femoral OP | 4.41 | (3.61, 5.40) | $<1.0 \times 10^{-40}$ | 4.63 | (3.56, 6.04) | $6.45 \times 10^{-30}$ | 8.68 | (5.88, 12.80) | $1.15 \times 10^{-27}$ |
| Lateral femoral OP | 4.89 | (3.34, 7.16) | $3.76 \times 10^{-16}$ | 8.02 | (5.26, 12.24) | $4.74 \times 10^{-22}$ | 13.57 | (8.04, 22.90) | $1.68 \times 10^{-22}$ |
| Medial tibial OP | 4.82 | (3.93, 5.93) | $<1.0 \times 10^{-40}$ | 6.42 | (4.99, 8.26) | $<1.0 \times 10^{-40}$ | 13.67 | (9.54, 19.60) | $<1.0 \times 10^{-40}$ |
| Lateral tibial OP | 3.23 | (2.66, 3.93) | $8.43 \times 10^{-32}$ | 4.14 | (3.20, 5.35) | $2.30 \times 10^{-27}$ | 6.86 | (4.65, 10.10) | $2.15 \times 10^{-22}$ |
| JSN | 2.64 | (2.13, 3.28) | $1.69 \times 10^{-18}$ | 3.77 | (2.85, 4.98) | $1.81 \times 10^{-20}$ | 5.77 | (3.80, 8.77) | $2.17 \times 10^{-16}$ |
| *Adjusted* |  |  |  |  |  |  |  |  |  |
| Any OP | 3.46 | (2.96, 4.04) | $<1.0 \times 10^{-40}$ | 4.40 | (3.57, 5.43) | $<1.0 \times 10^{-40}$ | 8.36 | (5.86, 11.91) | $7.64 \times 10^{-32}$ |
| OP at all locations | 4.33 | (2.03, 9.23) | $1.52 \times 10^{-04}$ | 4.03 | (1.68, 9.68) | 0.002 | 6.03 | (2.19, 16.59) | $5.00 \times 10^{-04}$ |
| Medial femoral OP | 4.04 | (3.29, 4.96) | $<1.0 \times 10^{-40}$ | 3.86 | (2.95, 5.06) | $1.01 \times 10^{-22}$ | 6.59 | (4.42, 9.82) | $2.11 \times 10^{-20}$ |
| Lateral femoral OP | 4.23 | (2.87, 6.24) | $3.51 \times 10^{-13}$ | 6.34 | (4.11, 9.76) | $5.45 \times 10^{-17}$ | 9.20 | (5.38, 15.72) | $4.94 \times 10^{-16}$ |
| Medial tibial OP | 4.36 | (3.53, 5.37) | $<1.0 \times 10^{-40}$ | 5.33 | (4.12, 6.91) | $7.63 \times 10^{-37}$ | 10.48 | (7.21, 15.23) | $6.48 \times 10^{-35}$ |
| Lateral tibial OP | 2.92 | (2.39, 3.56) | $8.07 \times 10^{-26}$ | 3.45 | (2.66, 4.49) | $1.97 \times 10^{-20}$ | 5.21 | (3.50, 7.75) | $4.03 \times 10^{-16}$ |
| JSN | 2.50 | (2.00, 3.12) | $6.44 \times 10^{-16}$ | 3.25 | (2.44, 4.33) | $7.60 \times 10^{-16}$ | 4.45 | (2.90, 6.82) | $7.26 \times 10^{-12}$ |

**Supplementary Table 8:**
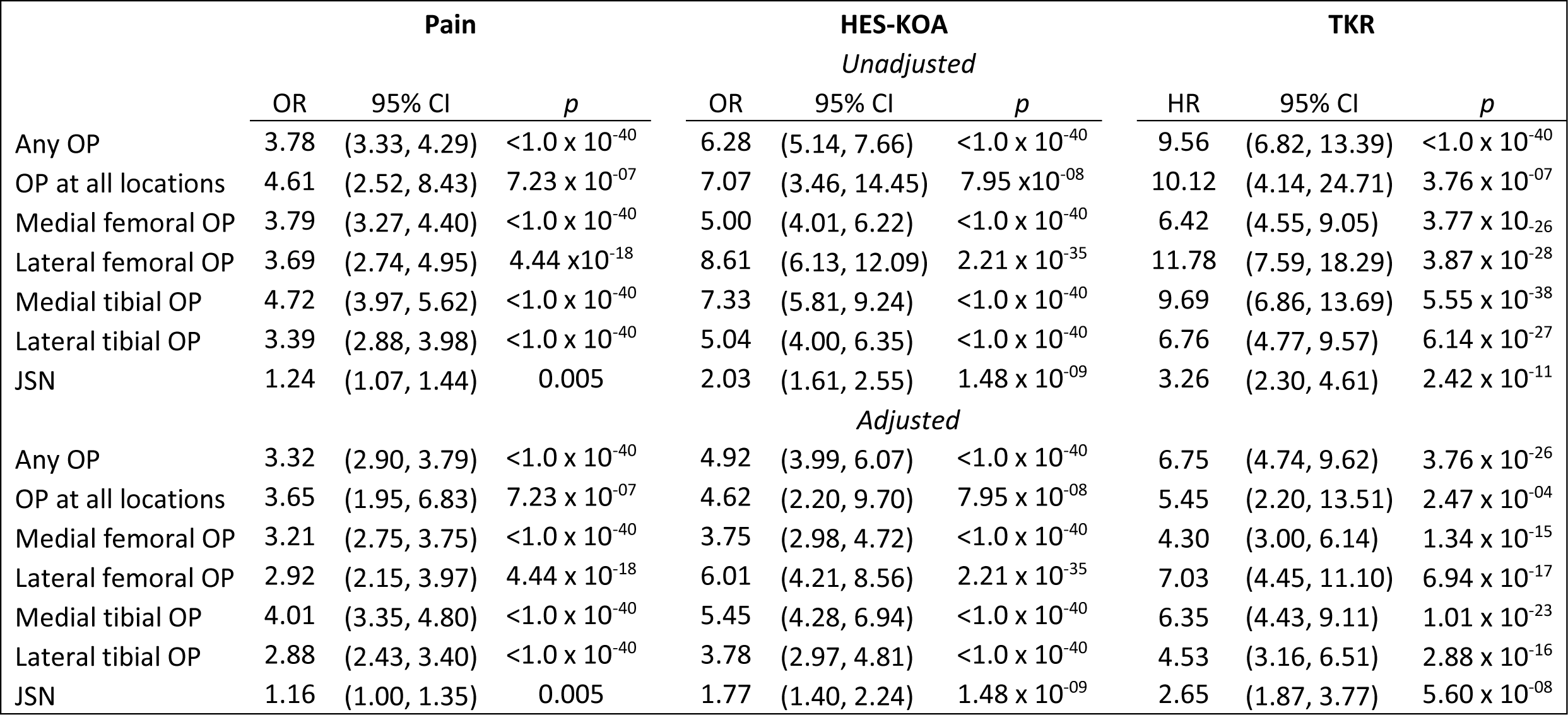
The associations of osteophytes and joint space narrowing (grades >1) with knee OA outcomes in females. Abbreviations: CI, Confidence Interval; HES-KOA, Knee osteoarthritis Based on Hospital Episodes Statistics; HR, Hazard Ratio; JSN, Joint Space Narrowing; OP, Osteophyte; OR Odds Ratio; TKR, Total Knee Replacement. Models were adjusted for age, height, weight and ethnic group. n=10,146.

**Supplementary Table 9:** Unadjusted associations of OP grades and JSN grades with knee OA outcomes in males and females combined. Abbreviations: CI, Confidence Interval; HES-KOA, Knee osteoarthritis based on Hospital Episodes Statistics; HR, Hazard Ratio; JSN, Joint Space Narrowing; OP, Osteophyte; OR Odds Ratio; TKR, Total Knee Replacement. *Denotes a sex-interaction term with p<0.05. n=19,595.

|  | Pain |  |  | HES KOA |  |  | TKR |  |  |
| --- | --- | --- | --- | --- | --- | --- | --- | --- | --- |
|  | OR | 95% CI | p | OR | 95% CI | p | HR | 95% CI | p |
| **Medial femur** |  |  |  |  |  |  |  |  |  |
| grade 1 | 2.93 | (2.49, 3.45) | $3.50 \times 10^{-38}$ | 2.86 | (2.23, 3.67) | $1.32 \times 10^{-16}$ | 3.98 | (2.66, 5.97) | $2.05 \times 10^{-11}$ |
| grade 2 | 5.07 | (4.12, 6.25) | $<1.0 \times 10^{-40}$ | 6.09 | (4.67, 7.94) | $<1.0 \times 10^{-40}$ | 9.73 | (6.71, 14.09) | $2.76 \times 10^{-33}$ |
| grade 3 | 6.40 | (4.91, 8.36) | $<1.0 \times 10^{-40}$ | 8.33 | (6.10, 11.38) | $<1.0 \times 10^{-40}$ | 13.38 | (8.88, 20.18) | $3.46 \times 10^{-35}$ |
| **Lateral femur** |  |  |  |  |  |  |  |  |  |
| grade 1 | 3.62 | (2.53, 5.17) | $1.65 \times 10^{-12}$ | 6.62 | (4.38, 10.02) | $3.44 \times 10^{-19}$ | 10.45 | (6.20, 17.62) | $1.30 \times 10^{-18}$ |
| grade 2 | 4.39 | (2.79, 6.89) | $1.31 \times 10^{-10}$ | 5.70 | (3.27, 9.91) | $7.44 \times 10^{-10}$ | 8.21 | (3.87, 17.42) | $4.07 \times 10^{-08}$ |
| grade 3 | 4.54 | (3.02, 6.83) | $3.59 \times 10^{-13}$ | 12.88 | (8.44, 19.65) | $2.09 \times 10^{-32}$ | 18.97 | (11.74, 30.66) | $3.05 \times 10^{-33}$ |
| **Medial tibia** |  |  |  |  |  |  |  |  |  |
| grade 1 | 4.63 | (3.96, 5.42) | $<1.0 \times 10^{-40}$ | 5.33 | (4.32, 6.58) | $<1.0 \times 10^{-40}$ | 8.56 | (6.27, 11.70) | $<1.0 \times 10^{-40}$ |
| grade 2 | 4.36 | (3.27, 5.83) | $2.05 \times 10^{-23}$ | 9.44 | (6.83, 13.06) | $<1.0 \times 10^{-40}$ | 16.19 | (10.80, 24.29) | $<1.0 \times 10^{-40}$ |
| grade 3 | 6.43 | (4.44, 9.32) | $8.65 \times 10^{-23}$ | 11.83 | (7.91, 17.69) | $2.77 \times 10^{-33}$ | 21.17 | (13.01, 34.42) | $9.02 \times 10^{-35}$ |
| **Lateral tibia** |  |  |  |  |  |  |  |  |  |
| grade 1 | 2.98 | (2.56, 3.48) | $<1.0 \times 10^{-40}$ | 3.43 | (2.75, 4.29) | $1.37 \times 10^{-27}$ | 4.56 | (3.21, 6.49) | $3.24 \times 10^{-17}$ |
| grade 2 | 3.67 | (2.81, 4.80) | $1.95 \times 10^{-21}$ | 6.51 | (4.72, 8.97) | $2.45 \times 10^{-30}$ | 10.43 | (6.82, 15.95) | $2.88 \times 10^{-27}$ |
| grade 3 | 4.41 | (3.31, 5.88) | $4.76 \times 10^{-24}$ | 6.80 | (4.81, 9.61) | $1.92 \times 10^{-27}$ | 11.81 | (7.59, 18.37) | $6.24 \times 10^{-28}$ |
| **JSN** |  |  |  |  |  |  |  |  |  |
| Grade 1 | 1.26 | (1.09, 1.45) | 0.002* | 1.69 | (1.36, 2.09) | $2.12 \times 10^{-06}$* | 2.50 | (1.78, 3.51) | $1.37 \times 10^{-07}$* |
| Grade 2 | 2.20 | (1.71, 2.84) | $9.71 \times 10^{-10}$* | 3.69 | (2.66, 5.12) | $4.92 \times 10^{-15}$* | 6.48 | (4.17, 10.06) | $1.01 \times 10^{-16}$ |
| Grade 3 | 5.25 | (3.41, 8.08) | $4.54 \times 10^{-14}$ | 9.63 | (6.00, 15.46) | $6.81 \times 10^{-21}$* | 17.59 | (10.40, 29.75) | $1.05 \times 10^{-26}$* |

**Supplementary Table 10:**
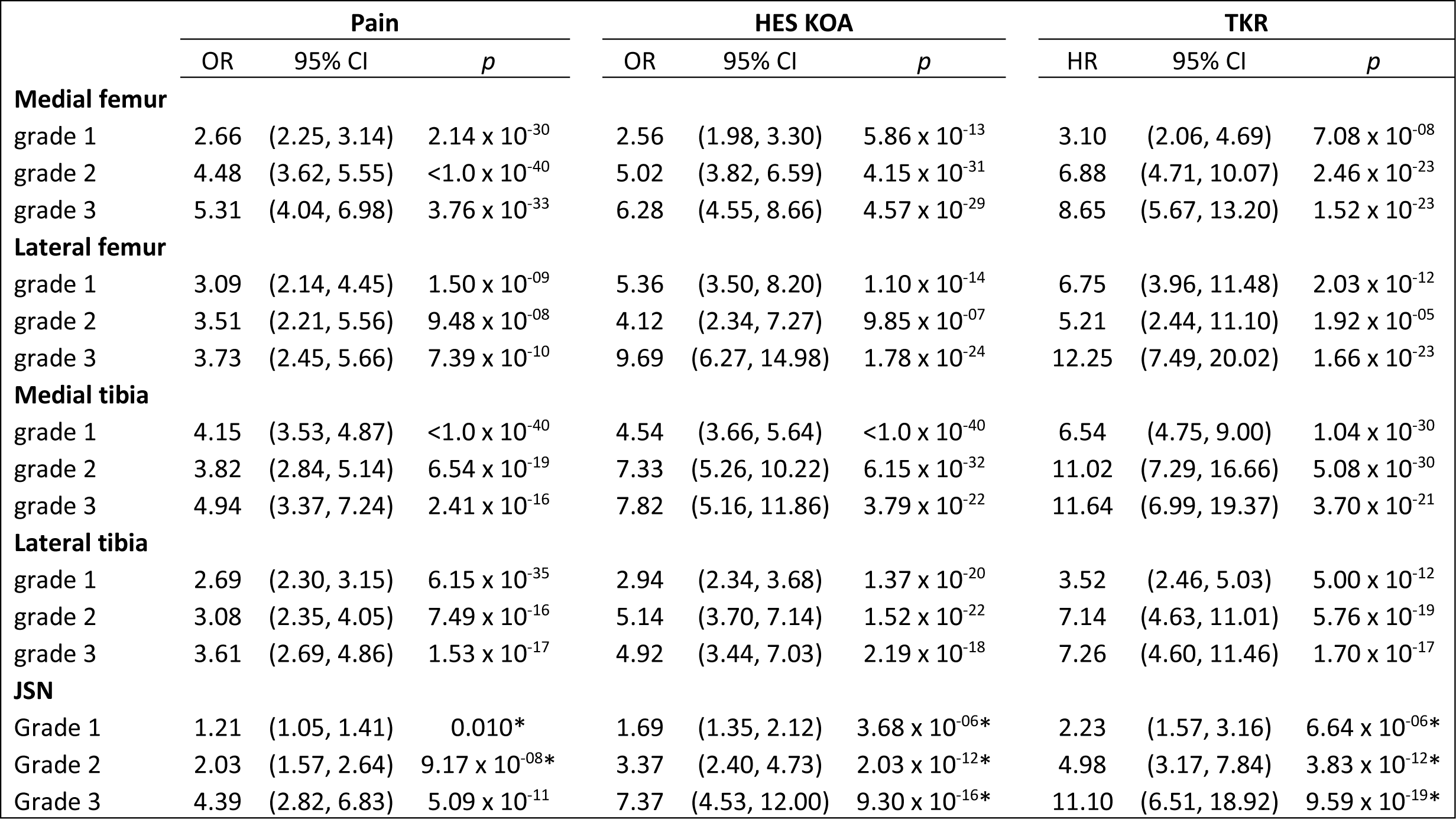
Adjusted associations of OP grades and JSN grades with knee OA outcomes in males and females combined. Abbreviations: CI, Confidence Interval; HES-KOA, Knee osteoarthritis based on Hospital Episodes Statistics; HR, Hazard Ratio; JSN, Joint Space Narrowing; OP, Osteophyte; OR Odds Ratio; TKR, Total Knee Replacement. *Denotes a sex-interaction term with p<0.05. Models were adjusted for age, sex, height, weight and ethnic group. n=19,595.

**Supplementary Table 11:** Unadjusted associations of OP grades and JSN grades with knee OA outcomes in males. Abbreviations: CI, Confidence Interval; HES-KOA, Knee osteoarthritis based on Hospital Episodes Statistics; HR, Hazard Ratio; JSN, Joint Space Narrowing; OP, Osteophyte; OR Odds Ratio; TKR, Total Knee Replacement. n=9,449.

|  | Pain |  |  | HES KOA |  |  | TKR |  |  |
| --- | --- | --- | --- | --- | --- | --- | --- | --- | --- |
|  | OR | 95% CI | *p* | OR | 95% CI | *p* | HR | 95% CI | *p* |
| **Medial femur** |  |  |  |  |  |  |  |  |  |
| grade 1 | 3.63 | (2.70, 4.89) | $1.79 \times 10^{-17}$ | 3.30 | (2.17, 5.01) | $2.11 \times 10^{-08}$ | 5.84 | (3.12, 10.91) | $3.16 \times 10^{-08}$ |
| grade 2 | 4.30 | (3.05, 6.06) | $9.05 \times 10^{-17}$ | 5.67 | (3.74, 8.59) | $2.75 \times 10^{-16}$ | 11.00 | (6.27, 19.31) | $6.54 \times 10^{-17}$ |
| grade 3 | 6.88 | (4.54, 10.43) | $9.24 \times 10^{-20}$ | 6.24 | (3.79, 10.26) | $5.55 \times 10^{-13}$ | 11.38 | (5.93, 21.85) | $2.58 \times 10^{-13}$ |
| **Lateral femur** |  |  |  |  |  |  |  |  |  |
| grade 1 | 4.58 | (2.22, 9.45) | $3.87 \times 10^{-05}$ | 7.02 | (3.11, 15.85) | $2.75 \times 10^{-06}$ | 18.38 | (8.08, 41.79) | $3.76 \times 10^{-12}$ |
| grade 2 | 4.86 | (2.33, 10.13) | $2.41 \times 10^{-05}$ | 5.04 | (2.04, 12.43) | $4.52 \times 10^{-04}$ | 6.26 | (1.55, 25.33) | 0.010 |
| grade 3 | 5.10 | (2.92, 8.92) | $1.13 \times 10^{-08}$ | 10.86 | (6.05, 19.49) | $1.35 \times 10^{-15}$ | 15.01 | (7.33, 30.76) | $1.36 \times 10^{-13}$ |
| **Medial tibia** |  |  |  |  |  |  |  |  |  |
| grade 1 | 4.71 | (3.62, 6.12) | $5.32 \times 10^{-31}$ | 5.08 | (3.63, 7.12) | $2.71 \times 10^{-21}$ | 10.80 | (6.75, 17.28) | $3.14 \times 10^{-23}$ |
| grade 2 | 3.82 | (2.56, 5.70) | $5.51 \times 10^{-11}$ | 9.07 | (5.90, 13.97) | $1.18 \times 10^{-23}$ | 17.38 | (10.03, 30.12) | $2.46 \times 10^{-24}$ |
| grade 3 | 7.90 | (4.73, 13.18) | $2.63 \times 10^{-15}$ | 7.88 | (4.41, 14.08) | $3.32 \times 10^{-12}$ | 19.39 | (9.74, 38.62) | $3.32 \times 10^{-17}$ |
| **Lateral tibia** |  |  |  |  |  |  |  |  |  |
| grade 1 | 2.72 | (2.11, 3.49) | $7.07 \times 10^{-15}$ | 2.94 | (2.07, 4.19) | $2.22 \times 10^{-09}$ | 4.39 | (2.50, 7.69) | $2.41 \times 10^{-07}$ |
| grade 2 | 3.48 | (2.28, 5.31) | $7.33 \times 10^{-09}$ | 5.83 | (3.56, 9.56) | $2.57 \times 10^{-12}$ | 10.53 | (5.49, 20.20) | $1.44 \times 10^{-12}$ |
| grade 3 | 5.10 | (3.37, 7.73) | $1.52 \times 10^{-14}$ | 6.95 | (4.29, 11.25) | $3.32 \times 10^{-15}$ | 11.40 | (6.11, 21.29) | $2.16 \times 10^{-14}$ |
| **JSN** |  |  |  |  |  |  |  |  |  |
| Grade 1 | 2.28 | (1.76, 2.93) | $2.35 \times 10^{-10}$ | 2.81 | (1.98, 3.99) | $6.22 \times 10^{-09}$ | 4.82 | (2.92, 7.96) | $7.72 \times 10^{-10}$ |
| Grade 2 | 3.44 | (2.08, 5.69) | $1.46 \times 10^{-06}$ | 9.22 | (5.45, 15.59) | $1.15 \times 10^{-16}$ | 11.14 | (5.42, 22.87) | $5.28 \times 10^{-11}$ |
| Grade 3 | 5.48 | (2.79, 10.78) | $8.13 \times 10^{-07}$ | 4.33 | (1.78, 10.51) | $1.21 \times 10^{-03}$ | 5.00 | (1.23, 20.25) | 0.024 |

**Supplementary Table 12:** Adjusted associations of OP grades and JSN grades with knee OA outcomes in males. Abbreviations: CI, Confidence Interval; HES-KOA, Knee osteoarthritis based on Hospital Episodes Statistics; HR, Hazard Ratio; JSN, Joint Space Narrowing; OP, Osteophyte; OR Odds Ratio; TKR, Total Knee Replacement. Models were adjusted for age, sex, height, weight and ethnic group. n=9,449.

|  | Pain |  |  | HES KOA |  |  | TKR |  |  |
| --- | --- | --- | --- | --- | --- | --- | --- | --- | --- |
|  | OR | 95% CI | *p* | OR | 95% CI | *p* | HR | 95% CI | *p* |
| *Categorical* |  |  |  |  |  |  |  |  |  |
| **Medial femur** |  |  |  |  |  |  |  |  |  |
| grade 1 | 3.35 | (2.48, 4.52) | $2.97 \times 10^{-15}$ | 2.86 | (1.87, 4.36) | $1.08 \times 10^{-06}$ | 4.83 | (2.58, 9.06) | $8.97 \times 10^{-07}$ |
| grade 2 | 4.00 | (2.82, 5.67) | $8.06 \times 10^{-15}$ | 4.80 | (3.14, 7.32) | $3.59 \times 10^{-13}$ | 8.54 | (4.84, 15.08) | $1.44 \times 10^{-13}$ |
| grade 3 | 6.06 | (3.97, 9.23) | $5.70 \times 10^{-17}$ | 4.76 | (2.86, 7.90) | $1.68 \times 10^{-09}$ | 7.24 | (3.70, 14.16) | $7.14 \times 10^{-09}$ |
| **Lateral femur** |  |  |  |  |  |  |  |  |  |
| grade 1 | 4.20 | (2.01, 8.76) | $1.29 \times 10^{-04}$ | 5.63 | (2.46, 12.90) | $4.32 \times 10^{-05}$ | 12.17 | (5.30, 27.90) | $3.63 \times 10^{-09}$ |
| grade 2 | 3.66 | (1.73, 7.76) | $6.88 \times 10^{-04}$ | 3.34 | (1.33, 8.39) | $1.03 \times 10^{-02}$ | 3.59 | (0.88, 14.71) | 0.076 |
| grade 3 | 4.61 | (2.62, 8.13) | $1.21 \times 10^{-07}$ | 9.37 | (5.17, 16.99) | $1.70 \times 10^{-13}$ | 11.39 | (5.53, 23.48) | $4.30 \times 10^{-11}$ |
| **Medial tibia** |  |  |  |  |  |  |  |  |  |
| grade 1 | 4.35 | (3.33, 5.68) | $2.29 \times 10^{-27}$ | 4.37 | (3.10, 6.14) | $2.60 \times 10^{-17}$ | 8.72 | (5.42, 14.03) | $4.62 \times 10^{-19}$ |
| grade 2 | 3.42 | (2.27, 5.14) | $3.35 \times 10^{-09}$ | 7.49 | (4.82, 11.62) | $2.78 \times 10^{-19}$ | 13.09 | (7.48, 22.91) | $2.21 \times 10^{-19}$ |
| grade 3 | 6.61 | (3.92, 11.14) | $1.34 \times 10^{-12}$ | 5.90 | (3.25, 10.69) | $5.00 \times 10^{-09}$ | 13.19 | (6.41, 27.15) | $2.48 \times 10^{-12}$ |
| **Lateral tibia** |  |  |  |  |  |  |  |  |  |
| grade 1 | 2.54 | (1.97, 3.28) | $6.42 \times 10^{-13}$ | 2.54 | (1.78, 3.64) | $3.10 \times 10^{-07}$ | 3.53 | (2.01, 6.21) | $1.18 \times 10^{-05}$ |
| grade 2 | 2.98 | (1.94, 4.58) | $6.16 \times 10^{-07}$ | 4.74 | (2.87, 7.84) | $1.29 \times 10^{-09}$ | 7.73 | (3.98, 15.00) | $1.50 \times 10^{-09}$ |
| grade 3 | 4.35 | (2.85, 6.65) | $1.09 \times 10^{-11}$ | 5.43 | (3.32, 8.89) | $1.72 \times 10^{-11}$ | 8.02 | (4.21, 15.28) | $2.54 \times 10^{-10}$ |
| **JSN** |  |  |  |  |  |  |  |  |  |
| Grade 1 | 2.17 | (1.68, 2.81) | $4.34 \times 10^{-09}$ | 2.46 | (1.72, 3.50) | $6.91 \times 10^{-07}$ | 3.78 | (2.28, 6.28) | $2.80 \times 10^{-07}$ |
| Grade 2 | 3.10 | (1.86, 5.19) | $1.59 \times 10^{-05}$ | 7.76 | (4.52, 13.31) | $9.89 \times 10^{-14}$ | 8.39 | (4.05, 17.39) | $1.37 \times 10^{-08}$ |
| Grade 3 | 5.20 | (2.62, 10.32) | $2.48 \times 10^{-06}$ | 3.68 | (1.50, 9.02) | 0.004 | 3.67 | (0.90, 14.92) | 0.071 |

**Supplementary Table 13:** Unadjusted associations of OP grades and JSN grades with knee OA outcomes in females. Abbreviations: CI, Confidence Interval; HES-KOA, Knee osteoarthritis based on Hospital Episodes Statistics; HR, Hazard Ratio; JSN, Joint Space Narrowing; OP, Osteophyte; OR Odds Ratio; TKR, Total Knee Replacement. n=10,146.

|  | Pain |  |  | HES KOA |  |  | TKR |  |  |
| --- | --- | --- | --- | --- | --- | --- | --- | --- | --- |
|  | OR | 95% CI | *p* | OR | 95% CI | *p* | HR | 95% CI | *p* |
| **Medial femur** |  |  |  |  |  |  |  |  |  |
| grade 1 | 2.74 | (2.25, 3.34) | $1.26 \times 10^{-23}$ | 3.04 | (2.22, 4.17) | $5.25 \times 10^{-12}$ | 3.34 | (1.96, 5.68) | $9.18 \times 10^{-06}$ |
| grade 2 | 5.67 | (4.35, 7.38) | $8.54 \times 10^{-38}$ | 6.98 | (4.93, 9.88) | $6.54 \times 10^{-28}$ | 9.10 | (5.55, 14.93) | $2.36 \times 10^{-18}$ |
| grade 3 | 6.15 | (4.34, 8.71) | $1.65 \times 10^{-24}$ | 10.99 | (7.34, 16.45) | $2.92 \times 10^{-31}$ | 15.04 | (8.83, 25.63) | $2.01 \times 10^{-23}$ |
| **Lateral femur** |  |  |  |  |  |  |  |  |  |
| grade 1 | 3.36 | (2.23, 5.08) | $8.04 \times 10^{-09}$ | 7.18 | (4.43, 11.66) | $1.45 \times 10^{-15}$ | 8.27 | (4.20, 16.30) | $1.02 \times 10^{-09}$ |
| grade 2 | 4.12 | (2.33, 7.31) | $1.23 \times 10^{-06}$ | 6.53 | (3.24, 13.18) | $1.64 \times 10^{-07}$ | 9.43 | (3.85, 23.07) | $9.02 \times 10^{-07}$ |
| grade 3 | 3.99 | (2.19, 7.26) | $6.06 \times 10^{-06}$ | 15.46 | (8.39, 28.50) | $1.65 \times 10^{-18}$ | 23.96 | (12.55, 45.76) | $6.44 \times 10^{-22}$ |
| **Medial tibia** |  |  |  |  |  |  |  |  |  |
| grade 1 | 4.62 | (3.79, 5.62) | $<1.0 \times 10^{-40}$ | 5.95 | (4.53, 7.82) | $1.11 \times 10^{-37}$ | 7.30 | (4.81, 11.10) | $1.19 \times 10^{-20}$ |
| grade 2 | 5.07 | (3.33, 7.72) | $4.24 \times 10^{-14}$ | 9.71 | (5.92, 15.91) | $1.90 \times 10^{-19}$ | 14.97 | (8.20, 27.34) | $1.24 \times 10^{-18}$ |
| grade 3 | 5.08 | (2.94, 8.78) | $5.52 \times 10^{-09}$ | 18.19 | (10.32, 32.06) | $1.14 \times 10^{-23}$ | 23.35 | (11.75, 46.42) | $2.56 \times 10^{-19}$ |
| **Lateral tibia** |  |  |  |  |  |  |  |  |  |
| grade 1 | 3.18 | (2.61, 3.86) | $5.77 \times 10^{-31}$ | 4.08 | (3.05, 5.44) | $1.43 \times 10^{-21}$ | 4.73 | (3.00, 7.47) | $2.52 \times 10^{-11}$ |
| grade 2 | 3.83 | (2.70, 5.41) | $3.63 \times 10^{-14}$ | 7.49 | (4.90, 11.44) | $1.19 \times 10^{-20}$ | 10.49 | (5.98, 18.39) | $2.45 \times 10^{-16}$ |
| grade 3 | 3.87 | (2.59, 5.77) | $3.47 \times 10^{-11}$ | 6.81 | (4.13, 11.22) | $5.50 \times 10^{-14}$ | 12.12 | (6.49, 22.66) | $5.30 \times 10^{-15}$ |
| **JSN** |  |  |  |  |  |  |  |  |  |
| Grade 1 | 1.00 | (0.84, 1.20) | 0.960 | 1.49 | (1.12, 1.97) | 0.005 | 1.84 | (1.15, 2.92) | 0.011 |
| Grade 2 | 1.90 | (1.41, 2.56) | $2.24 \times 10^{-05}$ | 2.67 | (1.71, 4.15) | $1.46 \times 10^{-05}$ | 5.38 | (3.06, 9.43) | $4.57 \times 10^{-09}$ |
| Grade 3 | 5.07 | (2.90, 8.87) | $1.28 \times 10^{-08}$ | 15.98 | (8.93, 28.60) | $9.93 \times 10^{-21}$ | 28.97 | (16.21, 51.80) | $6.81 \times 10^{-30}$ |

**Supplementary Table 14:** Adjusted associations of OP grades and JSN grades with knee OA outcomes in females. Abbreviations: CI, Confidence Interval; HES-KOA, Knee osteoarthritis based on Hospital Episodes Statistics; HR, Hazard Ratio; JSN, Joint Space Narrowing; OP, Osteophyte; OR Odds Ratio; TKR, Total Knee Replacement. Models were adjusted for age, height, weight and ethnic group. n=10,146.

|  | Pain |  |  | HES KOA |  |  | TKR |  |  |
| --- | --- | --- | --- | --- | --- | --- | --- | --- | --- |
|  | OR | 95% CI | *p* | OR | 95% CI | *p* | HR | 95% CI | *p* |
| **Medial femur** |  |  |  |  |  |  |  |  |  |
| grade 1 | 2.40 | (1.96, 2.93) | $2.51 \times 10^{-17}$ | 2.40 | (1.74, 1.74) | $1.03 \times 10^{-07}$ | 2.38 | (1.39, 4.08) | 0.002 |
| grade 2 | 4.74 | (3.61, 6.21) | $3.32 \times 10^{-29}$ | 5.08 | (3.55, 3.55) | $6.10 \times 10^{-19}$ | 5.75 | (3.46, 9.55) | $1.46 \times 10^{-11}$ |
| grade 3 | 4.77 | (3.33, 6.83) | $1.78 \times 10^{-17}$ | 7.60 | (5.00, 5.00) | $2.84 \times 10^{-21}$ | 9.50 | (5.48, 16.46) | $1.03 \times 10^{-15}$ |
| **Lateral femur** |  |  |  |  |  |  |  |  |  |
| grade 1 | 2.74 | (1.79, 4.19) | $3.13 \times 10^{-06}$ | 5.15 | (3.12, 3.12) | $1.30 \times 10^{-10}$ | 5.07 | (2.55, 10.09) | $3.82 \times 10^{-06}$ |
| grade 2 | 3.39 | (1.89, 6.09) | $4.43 \times 10^{-05}$ | 4.85 | (2.37, 2.37) | $1.55 \times 10^{-05}$ | 6.38 | (2.59, 15.70) | $5.45 \times 10^{-05}$ |
| grade 3 | 2.86 | (1.53, 5.34) | 0.001 | 9.80 | (5.14, 5.14) | $4.07 \times 10^{-12}$ | 12.29 | (6.22, 24.29) | $5.35 \times 10^{-13}$ |
| **Medial tibia** |  |  |  |  |  |  |  |  |  |
| grade 1 | 4.01 | (3.27, 4.91) | $<1.0 \times 10^{-40}$ | 4.63 | (3.50, 3.50) | $9.21 \times 10^{-27}$ | 5.19 | (3.38, 7.95) | $4.44 \times 10^{-14}$ |
| grade 2 | 4.34 | (2.82, 6.68) | $2.50 \times 10^{-11}$ | 7.15 | (4.31, 4.31) | $2.77 \times 10^{-14}$ | 9.11 | (4.93, 16.84) | $1.75 \times 10^{-12}$ |
| grade 3 | 3.52 | (1.99, 6.22) | $1.45 \times 10^{-05}$ | 10.49 | (5.79, 5.79) | $9.08 \times 10^{-15}$ | 10.61 | (5.17, 21.78) | $1.21 \times 10^{-10}$ |
| **Lateral tibia** |  |  |  |  |  |  |  |  |  |
| grade 1 | 2.77 | (2.27, 3.39) | $3.50 \times 10^{-23}$ | 3.20 | (2.38, 2.38) | $1.55 \times 10^{-14}$ | 3.41 | (2.14, 5.42) | $2.21 \times 10^{-07}$ |
| grade 2 | 3.14 | (2.20, 4.48) | $3.25 \times 10^{-10}$ | 5.45 | (3.53, 3.53) | $2.21 \times 10^{-14}$ | 6.76 | (3.81, 11.99) | $6.44 \times 10^{-11}$ |
| grade 3 | 3.05 | (2.01, 4.61) | $1.38 \times 10^{-07}$ | 4.41 | (2.62, 2.62) | $2.22 \times 10^{-08}$ | 6.65 | (3.48, 12.71) | $9.50 \times 10^{-09}$ |
| **JSN** |  |  |  |  |  |  |  |  |  |
| Grade 1 | 0.96 | (0.80, 1.15) | 0.665 | 1.37 | (1.03, 1.03) | 0.032 | 1.61 | (1.01, 2.57) | 0.045 |
| Grade 2 | 1.74 | (1.28, 2.36) | $3.61 \times 10^{-04}$ | 2.13 | (1.35, 1.35) | 0.001 | 3.85 | (2.18, 6.82) | $3.65 \times 10^{-06}$ |
| Grade 3 | 3.76 | (2.11, 6.70) | $7.28 \times 10^{-06}$ | 10.20 | (5.55, 5.55) | $7.32 \times 10^{-14}$ | 15.45 | (8.47, 28.16) | $4.17 \times 10^{-19}$ |

**Supplementary table 15:** Association of radiographic knee osteoarthritis (rKOA) grades with clinical outcomes, overall and stratified by sex. Abbreviations: CI, Confidence Interval; HES-KOA, Knee Osteoarthritis Based on Hospital Episodes Statistics; HR, Hazard Ratio; JSN, Joint Space Narrowing; OP, Osteophyte; OR Odds Ratio; TKR, Total Knee Replacement. *Denotes a sex-interaction term with p<0.05. Models were adjusted for age, sex, height, weight and ethnic group. Overall, n=19,595; males, n=9449; females n=10,146

| rKOA exposure | Unadjusted |  |  |  |  |  |  |  |  |
| --- | --- | --- | --- | --- | --- | --- | --- | --- | --- |
|  | Pain |  |  | HES KOA |  |  | TKR |  |  |
|  | OR | 95% CI | *p* | OR | 95% CI | *p* | HR | 95% CI | *p* |
| **All** |  |  |  |  |  |  |  |  |  |
| Grade 1 | 2.09 | (1.89, 2.31) | $<1.0 \times 10^{-40*}$ | 2.68 | (2.27, 3.15) | $3.29 \times 10^{-32}$ | 4.45 | (3.28, 6.03) | $5.37 \times 10^{-22}$ |
| Grade 2 | 3.87 | (3.29, 4.55) | $<1.0 \times 10^{-40}$ | 7.75 | (6.30, 9.52) | $<1.0 \times 10^{-40*}$ | 15.75 | (11.43, 21.71) | $<1.0 \times 10^{-40*}$ |
| Grades 3-4 | 8.62 | (6.63, 11.21) | $<1.0 \times 10^{-40}$ | 13.40 | (9.97, 18.01) | $<1.0 \times 10^{-40*}$ | 32.58 | (22.50, 47.17) | $<1.0 \times 10^{-40*}$ |
| **Male** |  |  |  |  |  |  |  |  |  |
| Grade 1 | 2.73 | (2.31, 3.21) | $4.10 \times 10^{-33}$ | 3.44 | (2.71, 4.37) | $3.91 \times 10^{-24}$ | 5.90 | (3.81, 9.13) | $1.83 \times 10^{-15}$ |
| Grade 2 | 4.53 | (3.43, 6.00) | $2.67 \times 10^{-26}$ | 11.51 | (8.45, 15.68) | $<1.0 \times 10^{-40}$ | 24.54 | (15.84, 38.02) | $<1.0 \times 10^{-40}$ |
| Grades 3-4 | 8.89 | (6.02, 13.13) | $5.21 \times 10^{-28}$ | 7.37 | (4.58, 11.87) | $2.07 \times 10^{-16}$ | 18.44 | (9.86, 34.47) | $6.79 \times 10^{-20}$ |
| **Female** |  |  |  |  |  |  |  |  |  |
| Grade 1 | 1.87 | (1.64, 2.13) | $5.75 \times 10^{-21}$ | 2.70 | (2.13, 3.40) | $7.80 \times 10^{-17}$ | 4.04 | (2.63, 6.21) | $2.10 \times 10^{-10}$ |
| Grade 2 | 3.66 | (2.99, 4.47) | $1.59 \times 10^{-36}$ | 7.09 | (5.31, 9.46) | $<1.0 \times 10^{-40}$ | 12.19 | (7.59, 19.58) | $4.17 \times 10^{-25}$ |
| Grades 3-4 | 8.48 | (5.94, 12.09) | $4.64 \times 10^{-32}$ | 23.09 | (15.62, 34.14) | $<1.0 \times 10^{-40}$ | 49.16 | (30.31, 79.71) | $<1.0 \times 10^{-40}$ |
| **Adjusted** |  |  |  |  |  |  |  |  |  |
| **All** |  |  |  |  |  |  |  |  |  |
| Grade 1 | 2.04 | (1.84, 2.26) | $<1.0 \times 10^{-40*}$ | 2.67 | (2.25, 3.16) | $1.47 \times 10^{-29}$ | 3.97 | (2.90, 5.42) | $5.64 \times 10^{-18}$ |
| Grade 2 | 3.65 | (3.08, 4.31) | $<1.0 \times 10^{-40}$ | 7.26 | (5.85, 9.00) | $<1.0 \times 10^{-40*}$ | 12.87 | (9.22, 17.96) | $<1.0 \times 10^{-40*}$ |
| Grades 3-4 | 7.08 | (5.41, 9.27) | $<1.0 \times 10^{-40}$ | 10.24 | (7.53, 13.93) | $<1.0 \times 10^{-40*}$ | 21.11 | (14.28, 31.19) | $<1.0 \times 10^{-40*}$ |
| **Male** |  |  |  |  |  |  |  |  |  |
| Grade 1 | 2.62 | (2.22, 3.10) | $1.20 \times 10^{-29}$ | 3.07 | (2.41, 3.91) | $1.46 \times 10^{-19}$ | 4.90 | (3.14, 7.63) | $2.33 \times 10^{-12}$ |
| Grade 2 | 4.34 | (3.27, 5.76) | $3.65 \times 10^{-24}$ | 10.06 | (7.35, 13.78) | $<1.0 \times 10^{-40}$ | 19.61 | (12.55, 30.65) | $<1.0 \times 10^{-40}$ |
| Grades 3-4 | 7.59 | (5.10, 11.29) | $1.65 \times 10^{-23}$ | 5.71 | (3.50, 9.30) | $2.65 \times 10^{-12}$ | 12.62 | (6.59, 24.17) | $2.13 \times 10^{-14}$ |
| **Female** |  |  |  |  |  |  |  |  |  |
| Grade 1 | 1.74 | (1.52, 1.99) | $3.61 \times 10^{-16}$ | 2.34 | (1.84, 2.96) | $2.51 \times 10^{-12}$ | 3.28 | (2.12, 5.07) | $9.14 \times 10^{-08}$ |
| Grade 2 | 3.22 | (2.61, 3.97) | $3.90 \times 10^{-28}$ | 5.58 | (4.14, 7.53) | $1.74 \times 10^{-29}$ | 8.75 | (5.39, 14.22) | $1.96 \times 10^{-18}$ |
| Grades 3-4 | 6.53 | (4.52, 9.43) | $1.66 \times 10^{-23}$ | 15.52 | (10.31, 23.35) | $<1.0 \times 10^{-40}$ | 28.63 | (17.09, 47.96) | $3.33 \times 10^{-37}$ |

**Supplementary Table 16:** Prevalence of height-adjusted radiographic knee osteoarthritis (rKOA) Radiographic knee osteoarthritis (rKOA) grades were generated based on a combination of osteophyte grades and joint space narrowing (JSN) grades. In the sensitivity analysis, the minimum joint space width (mJSW) of the medial compartment, which was used to define JSN grade, was normalised by the mean height of the population to account for height-related variations in mJSW.

|  | **Total**<br>N=19595 | **Male**<br>N=9449 | **Female**<br>N=10146 |
| --- | --- | --- | --- |
| **rKOA grade** |  |  |  |
| 0 | 14895 (76.01%) | 8366 (88.54%) | 6529 (64.35%) |
| 1 | 3312 (16.90%) | 787 (8.33%) | 2525 (24.89%) |
| 2 | 1128 (5.76%) | 198 (2.10%) | 930 (9.17%) |
| 3 | 182 (0.93%) | 69 (0.73%) | 113 (1.11%) |
| 4 | 78 (0.40%) | 29 (0.31%) | 49 (0.48%) |

**Supplementary Table 17:** Results of the sensitivity analysis looking at the association of height-adjusted rKOA grades with clinical outcomes. This table presents the regression results for the association of height-adjusted radiographic knee osteoarthritis (rKOA) grades with knee osteoarthritis outcomes. The outcomes evaluated include prolonged knee pain, hospital-diagnosed knee OA (HES-KOA), and subsequent total knee replacement (TKR). Analyses were adjusted for age, sex (in combined analysis), height, weight and ethnic category. *Denotes a sex-interaction term with p<0.05.

|  | Pain |  |  | HES-KOA |  |  | TKR |  |  |
| --- | --- | --- | --- | --- | --- | --- | --- | --- | --- |
|  | OR | lower CI | p | OR | lower CI | p | HR | lower CI | p |
| **All** |  |  |  |  |  |  |  |  |  |
| Grade 1 | 1.93 | (1.73, 2.14) | $2.05 \times 10^{-34*}$ | 2.69 | (2.26, 3.20) | $3.18 \times 10^{-29*}$ | 3.47 | (2.51, 4.80) | $4.61 \times 10^{-14*}$ |
| Grade 2 | 2.61 | (2.24, 3.03) | $4.09 \times 10^{-35*}$ | 5.08 | (4.09, 6.32) | $<1.0 \times 10^{-40*}$ | 9.19 | (6.51, 12.98) | $2.04 \times 10^{-36*}$ |
| Grades 3-4 | 7.40 | (5.72, 9.58) | $<1.0 \times 10^{-40}$ | 10.67 | (7.90, 14.39) | $<1.0 \times 10^{-40*}$ | 19.72 | (13.40, 29.02) | $<1.0 \times 10^{-40}$ |
| Grade $\geq 1$ | 2.28 | (2.08, 2.50) | $<1.0 \times 10^{-40*}$ | 3.62 | (3.12, 4.21) | $<1.0 \times 10^{-40*}$ | 5.86 | (4.47, 7.69) | $1.76 \times 10^{-37*}$ |
| **Males** |  |  |  |  |  |  |  |  |  |
| Grade 1 | 2.81 | (2.37, 3.33) | $1.95 \times 10^{-32}$ | 3.91 | (3.09, 4.95) | $6.95 \times 10^{-30}$ | 5.78 | (3.76, 8.88) | $1.34 \times 10^{-15}$ |
| Grade 2 | 4.40 | (3.27, 5.92) | $1.36 \times 10^{-22}$ | 9.18 | (6.57, 12.85) | $2.22 \times 10^{-38}$ | 18.63 | (11.74, 29.56) | $2.33 \times 10^{-35}$ |
| Grades 3-4 | 7.47 | (4.95, 11.26) | $8.72 \times 10^{-22}$ | 5.26 | (3.14, 8.81) | $3.05 \times 10^{-10}$ | 12.06 | (6.16, 23.61) | $3.74 \times 10^{-13}$ |
| Grade $\geq 1$ | 3.37 | (2.91, 3.90) | $<1.0 \times 10^{-40}$ | 4.88 | (3.99, 5.96) | $<1.0 \times 10^{-40}$ | 8.67 | (6.06, 12.41) | $3.10 \times 10^{-32}$ |
| **Females** |  |  |  |  |  |  |  |  |  |
| Grade 1 | 1.56 | (1.36, 1.78) | $5.54 \times 10^{-11}$ | 1.85 | (1.45, 2.37) | $7.97 \times 10^{-07}$ | 2.02 | (1.27, 3.22) | 0.003 |
| Grade 2 | 2.11 | (1.76, 2.53) | $5.96 \times 10^{-16}$ | 3.40 | (2.55, 4.53) | $9.29 \times 10^{-17}$ | 5.14 | (3.19, 8.29) | $1.94 \times 10^{-11}$ |
| Grades 3-4 | 6.90 | (4.95, 9.63) | $5.65 \times 10^{-30}$ | 13.93 | (9.49, 20.45) | $<1.0 \times 10^{-40}$ | 21.96 | (13.15, 36.66) | $3.36 \times 10^{-32}$ |
| Grade $\geq 1$ | 1.826 | (1.62, 2.05) | $9.71 \times 10^{-24}$ | 2.63 | (2.13, 3.25) | $2.11 \times 10^{-19}$ | 3.66 | (2.50, 5.37) | $2.85 \times 10^{-11}$ |

## Supplementary Methods

### Assessment of covariates

The selection of covariates was predetermined based on existing literature associating these variables with knee osteoarthritis risk. Height and weight measurements were taken at the time of the DXA scan, following standardized procedures, while age and sex were self-reported during recruitment. Participants self-reported their ethnicity, which was then categorized into groups including White, Black, Asian, Chinese, Mixed-Heritage and Other.

### Generation of a rKOA osteophyte grades

In this study, osteophyte area cut-offs were generated using a combination of manually graded osteophyte grades and shading of osteophyte area. Binary variables were first created for Grade 2 and Grade 3 osteophytes in both the medial and lateral femur regions. These variables were assigned a value of 0 if the manually graded osteophyte was below the specified grade and a value of 1 if the manually graded osteophyte matched the specified grade. Optimal cut-points for osteophyte area were then estimated using the Youden Index method, which determines the optimal threshold for classification based on a continuous predictor variable, by maximizing the sum of sensitivity and specificity. The reference variable was the binary outcome variable, while the classification variable was the continuous osteophyte area measurement. The empirical optimal cut-points for Grade 2 and Grade 3 osteophytes were as follows: medial femur (grade 2: 14.72 mm^2^, grade 3: 25.78 mm), lateral femur (14.03 mm^2^, 24.62 mm^2^), medial tibia (12.20 mm^2^, 20.28 mm^2^), and lateral tibia (10.53 mm^2^, 18.65 mm^2^). Based on these cut-offs, osteophyte grades were determined accordingly:

| Grade | Medial Femur |  | Lateral Femur |  | Medial Tibia |  | Lateral Tibia |  |
| --- | --- | --- | --- | --- | --- | --- | --- | --- |
|  | lower | upper | lower | upper | lower | upper | lower | upper |
| 1 | $>0 \text{ mm}^2$ | $\leq 15 \text{ mm}^2$ | $>0 \text{ mm}^2$ | $\leq 14 \text{ mm}^2$ | $>0 \text{ mm}^2$ | $\leq 12 \text{ mm}^2$ | $>0 \text{ mm}^2$ | $\leq 11 \text{ mm}^2$ |
| 2 | $\geq 15 \text{ mm}^2$ | $< 26 \text{ mm}^2$ | $\geq 14 \text{ mm}^2$ | $< 25 \text{ mm}^2$ | $\geq 12 \text{ mm}^2$ | $\leq 20 \text{ mm}^2$ | $\geq 11 \text{ mm}^2$ | $\leq 19 \text{ mm}^2$ |
| 3 | $\geq 26 \text{ mm}^2$ | | $\geq 25 \text{ mm}^2$ | | $\geq 20 \text{ mm}^2$ | | $\geq 19 \text{ mm}^2$ | |

These grades were then multiplied by 0.5, summed (with a maximum value of 6), and added to the JSN grade (which has a maximum value of 3) to derive a combined score, with a maximum value of 9. Subsequently, the overall rKOA grade was determined based on this combined score, as detailed in the main text.

### Sensitivity analysis

Height normalisation of medial joint space width (mJSW) was performed to mitigate potential confounding effects of participant height on mJSW measurements. Initially, a linear regression analysis was conducted using height as the independent variable and mJSW as the dependent variable. This regression provided a beta coefficient representing the change in mJSW per unit change in height. Subsequently, for each participant, the difference between their height and the mean height of the study population was calculated. This difference was multiplied by the beta coefficient to derive an inflation factor, which served to adjust each individual’s mJSW measurement based on their height deviation from the mean. Adjustments were made by adding the inflation factor for individuals taller than the mean height and subtracting it for those shorter. These normalised mJSW values were then used to redefine Joint Space Narrowing (JSN) grades as before (JSN grade 0 for mJSW >3mm, grade 1 for mJSW >2.5mm and <3mm, grade 2 for mJSW >2mm and <2.5mm, and grade 3 for mJSW <2mm). The revised JSN grades were integrated with osteophyte grades to compute overall radiographic knee osteoarthritis (rKOA) grade, consistent with the methodology detailed in the main text. This approach ensured that mJSW measurements accurately reflected joint space narrowing independent of height variations.

